# Inference of causal relationships based on the genetics of cardiometabolic traits and conditions unique to females in >50,000 participants

**DOI:** 10.1101/2022.02.02.22269844

**Authors:** Brenda Xiao, Digna R. Velez Edwards, Anastasia Lucas, Theodore Drivas, Kathryn Gray, Brendan Keating, Chunhua Weng, Gail P. Jarvik, Hakon Hakonarson, Leah Kottyan, Noemie Elhadad, Regeneron Genetics Center, Wei-Qi Wei, Yuan Luo, Dokyoon Kim, Marylyn Ritchie, Shefali Setia Verma

**Affiliations:** University of Pennsylvania; Division of Quantitative Sciences, Department of Obstetrics and Gynecology, Vanderbilt University Medical Center; Department of Genetics, University of Pennsylvania; Brigham and Women’s Hospital Department of Obstetrics and Gynecology; Department of Surgery, University of Pennsylvania; Department of Biomedical Informatics, Columbia University; Departments of Medicine (Medical Genetics) and Genome Sciences, University of Washington Medical Center; Center for Applied Genomics, Children’s Hospital of Philadelphia; Center for Autoimmune Genomics and Etiology and Division of Allergy & Immunology, Cincinnati Children’s Hospital Medical Center, Department of Pediatrics, University of Cincinnati; Department of Biomedical Informatics, Vanderbilt University Medical Center; Department of Preventive Medicine, Feinberg School of Medicine, Northwestern University; Department of Biostatistics and Epidemiology, University of Pennsylvania; Department of Pathology and Laboratory Medicine, University of Pennsylvania

## Abstract

**Background:** Cardiometabolic diseases are highly comorbid and associated with poor health outcomes. However, the investigation of the relationship between the genetic predisposition to cardiometabolic diseases with the risk of conditions unique to females such as breast cancer, endometriosis and pregnancy-related complications is highly understudied. This study aimed to estimate the cross-trait genetic overlap and influence of genetic burden of cardiometabolic traits on health conditions unique to females.

**Methods:** We obtained data for female participants in the Penn Medicine BioBank (PMBB; 21,837 samples) and the electronic MEdical Records and GEnomics (eMERGE; 49,171 samples) network. We examined the relationship between four cardiometabolic phenotypes (body mass index (BMI), coronary artery disease (CAD), type 2 diabetes (T2D) and hypertension (through blood pressure measurements)) and 23 female health conditions by performing four analyses: 1) Cross-trait genetic correlation analyses to compare genetic architecture. 2) Polygenic risk scores (PRS)-based association tests to characterize shared genetic effects on disease risk. 3) Mendelian randomization (MR) for significant associations to assess cross-trait causal relationships. 4) Chronology analyses to visualize the timeline of events unique to groups of females with high and low genetic burden for cardiometabolic traits and highlight the disease prevalence in risk groups by age.

**Results:** We observed high genetic correlation among cardiometabolic and female health conditions. PRS meta-analysis identified 29 significant associations reflecting potential shared biology among common cardiometabolic phenotypes and female health conditions. Significant associations include PRS_BMI_ with endometrial cancer and polycystic ovarian syndrome (PCOS), PRS_CAD_ with breast cancer, and the PRS_T2D_ with gestational diabetes and PCOS. Mendelian randomization provided additional evidence of independent causal effects between T2D and gestational diabetes and CAD and with breast cancer. Our results reflected inverse association between PRS_CAD_ and breast cancer. Lastly, as visualized from chronology analyses, individuals with high PRS are also more likely to develop conditions such as PCOS and gestational hypertension at earlier ages.

**Conclusions:** Polygenic susceptibility to cardiometabolic traits is associated with conditions unique to females. Several of these associations are likely to result from the complex pathophysiology of cardiometabolic risk, and others may reflect potential pleiotropic effects that go beyond cardiometabolic health in females.

## Introduction

Cardiometabolic diseases such as coronary artery disease (CAD), obesity, hypertension, and type 2 diabetes (T2D) are profoundly prevalent and among the leading causes of death in the world^5,6,7^. Cardiometabolic conditions are highly comorbid and considered as risk factors for sequelae of many diseases, such as depression, anxiety, chronic obstructive pulmonary disease (COPD), and cancer, to name a few^8, 9^. These conditions affect female individuals disproportionately because they are also linked to disorders of the reproductive system and adverse outcomes of pregnancy and childbirth such as preeclampsia, gestational diabetes, stillbirth, and pregnancy loss^10, 11^. Many studies suggest that the pathophysiology of cardiometabolic diseases affects males and females differently^14^. There are multiple shreds of evidence supporting the relationship between female health conditions and cardiometabolic diseases. For instance, people who develop preeclampsia during pregnancy are more likely to develop cardiovascular diseases and hypertension after pregnancy^15, 16^. Additionally, obesity and polycystic ovarian syndrome (PCOS) are closely linked conditions, and people with PCOS are at high risk of developing T2D^17–19^. People with endometriosis are also at high risk for developing oncological cancers and cardiovascular diseases such as myocardial infarction and ischemic heart disease^20, 21^. However, the relationship between female health and cardiometabolic phenotypes, particularly the potential for any shared genetic burden between them, is still highly understudied. The investigation of shared genetic burden could lead to identifying obesity and cardiometabolic cluster-related risk factors associated with female health conditions to modify standard screening practices in people at high risk for many diseases.

Genome-wide association studies (GWAS) in the past couple of decades have exposed common cross-trait connections. Disease traits such as type 2 diabetes, obesity, sleep apnea, hypertension, Alzheimer’s diseases, and many cancer types have been shown to share genetic etiology^1–4^. GWAS have identified >1000 loci with a likely impact on cardiometabolic phenotypes, but the effect size of any single variant is generally tiny. Many methods have identified relationships between different phenotypes and traits by calculating their genetic correlation from GWAS effect sizes. In addition, to more accurately estimate an individual’s overall risk for disease, researchers have used GWAS to calculate polygenic risk scores (PRS), which sum up the effect of common single nucleotide polymorphisms (SNPs) throughout the genome into a single score of overall genetic burden for a phenotype^27^. PRS have been shown to predict the risk of many cardiometabolic diseases and its comorbidities^28, 29^.

Our approach to investigating the impact of genetic burden of cardiometabolic traits in female health conditions is to measure the genetic correlation and association of cardiometabolic PRS with EHR-derived phenotypes. Many prior studies have successfully used PRS as the genetic risk factor for identifying links between overall genetic risk for one phenotype to other phenotypes^31, 32^. PRS-based association tests make fewer assumptions and have the benefit that they are based on unvarying risk factors (i.e., inherited genetic burden). We hypothesize that the genetic burden for cardiometabolic phenotypes could further explain the phenotypic variance in health conditions unique to females. This present study obtained genotyped data and a wide range of female health conditions from the Penn Medicine BioBank (PMBB) and the Electronic Medical Records and Genomics (eMERGE) Network. We measured pairwise genetic correlation between cardiometabolic phenotypes and female health conditions to identify the strength of shared genetic factors. The association of PRS with multiple selected phenotypes is further used to estimate the significance of associations between cardiometabolic genetic burden and female health conditions. We additionally evaluated the causal relationship between significant associations by using Mendelian randomization (MR) approaches. Lastly, electronic health record (EHR) data provides the opportunity to map female health by considering the disease prevalence by age. We generated a chronological map of diseases in participants in high and low PRS groups to understand the prevalence of diseases in the two risk groups at different ages.

## Methods

The authors have individual level access to genotype and medical record data from the Penn Medicine BioBank and the Electronic Medical Records and Genomics network datasets.

### Study Populations

#### Penn Medicine BioBank

The Penn Medicine BioBank (PMBB) is a University of Pennsylvania academic biobank which recruits patient-participants from the University of Pennsylvania Health System around the greater Philadelphia area in the United States. PMBB links an individual’s genotype data with detailed electronic health record (EHR) information. Currently, PMBB consists of genome-wide genotyped and whole-exome sequenced data on ∼45,000 samples. PMBB is a diverse cohort, with over 25% of participants of African ancestry. PMBB genotyped data is imputed to TOPMED Reference panel using the Michigan Imputation server. We included 21,837 female participants from PMBB in this study (Table 1). The stratified analyses in this study only included participants of European and African ancestry and excluded those from Asian and Hispanic ancestries due to the limiting sample size.

**Table 1:** Sample size in PMBB and eMERGE datasets, for overall and stratified by ancestry analyses.

| Phenotype | PMBB Sample Size<br>(N Cases) |  |  | eMERGE Sample Size<br>(N Cases) |  |  |
| --- | --- | --- | --- | --- | --- | --- |
|  | All | EUR | AFR | All | EUR | AFR |
| *Cardiometabolic Phenotypes* |  |  |  |  |  |  |
| BMI | 20,209 | 12,344 | 6,744 | 39,403 | 31,875 | 5,250 |
| CAD | 21,837<br>(3,002) | 13,515<br>(1,956) | 7,039<br>(955) | 49,171<br>(9,597) | 37,003<br>(7,524) | 8,308<br>(1,308) |
| DBP | 21,612 | 13,343 | 6,994 | NA | NA | NA |
| Hypertension | 21,837<br>(10,278) | 13,515<br>(5,640) | 7,039<br>(4,286) | 49,171<br>(27,685) | 37,003<br>(20,820) | 8,308<br>(4,695) |
| T2D | 21,837<br>(4,388) | 13,515<br>(1,969) | 7,039<br>(2,221) | 49,171<br>(12,403) | 37,003<br>(8,311) | 8,308<br>(2,725) |
| *Women's Health Phenotypes* |  |  |  |  |  |  |
| Breast cancer | 21,837<br>(1,621) | 13,515<br>(1,621) | 7,039<br>(415) | 49,171<br>(4,148) | 37,003<br>(3,532) | 8,308<br>(408) |
| Cervical cancer | 21,837<br>(105) | 13,515<br>(53) | 7,039<br>(50) | 49,171<br>(332) | 37,003<br>(252) | 8,308<br>(52) |
| Ectopic pregnancy | 2,808<br>(1,779) | 1,201<br>(827) | 1,319<br>(757) | 3,078<br>(332) | 1,975<br>(521) | 641<br>(153) |
| Endometrial cancer | 21,837<br>(286) | 13,515<br>(183) | 7,039<br>(90) | 49,171<br>(771) | 37,003<br>(653) | 8,308<br>(74) |
| Endometriosis | 21,837<br>(701) | 13,515<br>(320) | 7,039<br>(340) | 49,171<br>(2,314) | 37,003<br>(1,805) | 8,308<br>(330) |
| Excessive fetal growth | 693<br>(85) | 293<br>(37) | 329<br>(42) | 2,433<br>(627) | 1,618<br>(377) | 437<br>(150) |
| Gestational diabetes | 2,655<br>(523) | 1,111<br>(193) | 1,262<br>(248) | 3,174<br>(762) | 2,005<br>(414) | 719<br>(244) |
| Gestational hypertension | 2,666<br>(631) | 1,118<br>(256) | 1,275<br>(324) | 3,135<br>(703) | 1,980<br>(383) | 711<br>(235) |
| Intrauterine death | 685<br>(57) | 289<br>(23) | 324<br>(27) | 2,156<br>(64) | 1,438<br>(38) | 358<br>(17) |
| Miscarriage | 2,934<br>(389) | 1,273<br>(199) | 1,360<br>(151) | 3,568<br>(823) | 2,323<br>(547) | 740<br>(182) |
| Ovarian cancer | 21,837<br>(305) | 13,515<br>(191) | 7,039<br>(89) | 49,171<br>(1,589) | 37,003<br>(1,324) | 8,308<br>(177) |
| Placenta<br>abruption/previa | 1,192<br>(396) | 482<br>(157) | 586<br>(195) | 2,674<br>(997) | 1,697<br>(616) | 583<br>(240) |
| Polycystic<br>ovarian<br>syndrome | 21,837<br>(736) | 13,515<br>(387) | 7,039<br>(272) | 49,171<br>(1,006) | 37,003<br>(690) | 8,308<br>(200) |
| Poor fetal<br>growth | 792<br>(202) | 325<br>(78) | 388<br>(107) | 2,209<br>(167) | 1,478<br>(102) | 360<br>(21) |
| Postpartum<br>depression | 924<br>(384) | 374<br>(136) | 466<br>(226) | 2,417<br>(555) | 1,579<br>(319) | 457<br>(157) |
| Postpartum<br>hemorrhage | 846<br>(283) | 350<br>(108) | 408<br>(146) | 2,320<br>(401) | 1,544<br>(250) | 408<br>(104) |
| Preeclampsia | 2,631<br>(452) | 1,100<br>(149) | 1,264<br>(272) | 3,132<br>(702) | 1,974<br>(377) | 713<br>(13) |
| Preterm birth | 687<br>(66) | 284<br>(21) | 332<br>(40) | 2,144<br>(57) | 1,433<br>(30) | 350<br>(13) |
| Stillbirth | 649<br>(17) | 270<br>(3) | 311<br>(12) | 2,123<br>(8) | 1,417<br>(6) | 347<br>(0) |
| Uterine cancer | 21,837<br>(113) | 13,515<br>(66) | 7,039<br>(43) | 49,171<br>(418) | 37,003<br>(340) | 8,308<br>(58) |
| Uterine fibroid | 21,837<br>(1,570) | 13,515<br>(516) | 7,039<br>(984) | 49,171<br>(5,711) | 37,003<br>(3,912) | 8,308<br>(1,347) |
| Vaginal cancer | 21,837<br>(23) | 13,515<br>(13) | 7,039<br>(9) | 49,171<br>(109) | 37,003<br>(86) | 8,308<br>(17) |
| Vulvar cancer | 21,837<br>(41) | 13,515<br>(25) | 7,039<br>(14) | 49,171<br>(120) | 37,003<br>(92) | 8,308<br>(19) |

#### eMERGE

The Electronic Medical Records and Genomics (eMERGE) Network is a nationwide consortium containing individuals with genome-wide genotyped data linked to EHRs from several health systems across the United States, with most participants from the Geisinger Health System and Vanderbilt University. eMERGE data is imputed to Haplotype Reference Consortium panel using Michigan Imputation Server. Like PMBB, the eMERGE cohort is diverse across ancestries and ages. This study was performed on 49,171 female patients in eMERGE born after 2001 (Table 1).

### Genome-wide association studies for cardiometabolic phenotypes

Most genetic correlation and PRS calculation methods require effect sizes of variants on the phenotypes determined through large GWAS. Given the diverse nature of our study population, we obtained the largest publicly available multi-ancestry GWAS summary statistics for six cardiometabolic phenotypes: obesity (measured through body mass index (BMI), CAD, hypertension (measured through diastolic blood pressure (DBP), systolic blood pressure (SBP), and pulse pressure (PP)), and T2D. The summary statistics used for each phenotype and its respective study is referenced in Table 2.

**Table 2:** GWAS summary statistics datasets that are used for the calculation of genetic correlations and polygenic risk scores and the number of SNPs used from each GWAS to calculate the corresponding PRS.

| Phenotype | Ancestries Included | Source | Sample Size (Number of Cases) | PMID | N SNPs included in PRS |
| --- | --- | --- | --- | --- | --- |
| Type 2 Diabetes | EUR, AFR, EAS, SAS, HIS | Vujkovic et al., Nat Gen, 2020, MVP | 1,407,282<br>(228,499) | 32541925 | PMBB: 1,023,697<br>eMERGE: 716,330 |
| Body Mass Index | EUR, AFR, EAS, SAS, HIS | Justice et al., Nat Com, 2017, GIANT | 241,258 | 28443625 | PMBB: 885,143<br>eMERGE: 614,668 |
| Hypertension (DBP, SBP, PP) | EUR, AFR, EAS, SAS, HIS | Giri et al., Nat Gen, 2019, MVP | 318,891 | 30578418 | PMBB: 1,024,567<br>eMERGE: 715,471 |
| Coronary Artery Disease | EUR, EAS, SAS, HIS, AFR | van der Harst et al., Circ Res, 2018, CARDIoGRAMplusC4D + UKBB | 547,261 (122,733) | 29212778 | PMBB: 981,480<br>eMERGE: 681,029 |

\*\*DBP= Diastolic Blood Pressure; SBP= Systolic Blood Pressure; PP= Pulse Pressure; EUR= European ancestry; EAS= East Asian ancestry; SAS= South Asian ancestry; HIS= Hispanic ancestry; AFR= African ancestry; MVP= Million Veterans Program; GIANT=The Genetic Investigation of ANthropometric Traits; UKB= UK BioBank

### Genome-wide association for female health conditions

Large multi-ancestry GWAS are not available for most female health conditions evaluated in this study. Thus, we used PLINK version 1.90 to conduct GWAS for the female health conditions in PMBB and eMERGE datasets^33^. We filtered the variants in the PMBB imputed datasets to include only those with imputation quality R^2^ > 0.3 and minor allele frequency > 0.01. We then meta-analyzed the GWAS from our two cohorts in PLINK.

### Genetic correlation calculation

We calculated pairwise genetic correlation between cardiometabolic phenotypes and female health conditions using LD score regression (LDSC), with the large publicly available cardiometabolic GWAS and the meta-analyzed GWAS for female health conditions from PMBB and eMERGE as input GWAS^34^. LDSC accounts for linkage disequilibrium (LD) among SNPs by using an external reference panel that should also match the ancestry distribution of the GWAS. We generated a multi-ancestry LD reference panel using the HapMap3 SNPs (∼1M common variants) from the entire 1000 Genomes population.

### Polygenic Risk Scores (PRS)

PRS calculated using GWAS performed in one ancestry group tend to perform poorly in individuals from different ancestries^27, 35, 36^. To accurately calculate a PRS for our diverse target datasets (PMBB and eMERGE), we calculated a PRS for the six different cardiometabolic phenotypes using the large publicly available multi-ancestry GWAS used in the genetic correlation analyses. Weights for each SNP included in the PRS were calculated using PRS-CS (version from Apr 24, 2020)^38^. Like LDSC, PRS-CS requires a reference panel that matches the ancestry distribution of the target dataset, and we generated a similar multi-ancestry LD reference panel using the HapMap SNPs from 1000 Genomes. We used PLINK(v1.90) to identify LD blocks and calculate the LD between the SNPs in each block. For PRS-CS, the global shrinkage parameter phi was fixed to 0.01, and default values were selected for all other parameters. PRS was then calculated using these weights through PLINK. Only the SNPs in the target dataset, summary statistics, and LD reference panel were included in the PRS. The number of SNPs used for PRS calculation is listed in Table 2. Scores were then normalized to obtain meaningful beta coefficients.

To evaluate the power of PRS, we tested the performance of each PRS on predicting the primary phenotype of the summary statistics. We could not obtain any blood pressure quantitative measurements for participants from eMERGE, and PP and SBP measurements were not curated in PMBB. Therefore, we evaluated the performance of the PRS from blood pressure traits (SP, DBP, PP) in eMERGE on hypertension case-control phenotype and in PMBB by predicting DBP or hypertension (for SBP and PP) as outcomes. We constructed logistic regression models for binary phenotypes (CAD, T2D, and hypertension), and evaluated PRS performance based on the area under the receiver-operator curve (AUC) using R’s *pROC* package. Similarly, we constructed linear regression models for continuous phenotypes (BMI and DBP) and evaluated them based on the R^2^ using R’s *glm* function. The regression models used birth year and the first five principal components (PCs) as covariates. We tested PRS performance in all individuals as well as in only European and African ancestry individuals.

### Phenotype data

Cases and controls for each phenotype were defined using International Classification of Disease (ICD) diagnosis codes. Participants were coded as cases for a phenotype if they had at least one occurrence of the corresponding ICD codes. For pregnancy-related phenotypes, participants were only considered as controls for a phenotype if they had at least one occurrence of a pregnancy-related ICD code and no occurrences of codes for certain complications during pregnancy (such as miscarriage) or any of the case ICD codes. Participants were counted as controls for cardiometabolic and all other female health conditions if they did not have any of the case ICD codes. The complete list of all ICD codes used to include or exclude participants as cases and controls can be found in Supplementary Table 1. Using these definitions, we determined the sample size for each phenotype in eMERGE and PMBB (Table 1).

### PRS association analyses

We tested the association of each cardiometabolic PRS and female health conditions by fitting separate logistic regression models, adjusting the models by birth year and the first five PCs. We conducted this analysis in all participants as well as ancestry-stratified analyses for individuals of European and African ancestry. To account for biases from multiple hypothesis testing, we determined if these associations passed an FDR-significance threshold of 0.05, using the number of female health conditions (23) as the number of hypotheses tested. The logistic regressions were performed using R’s *glm* function, and results from PMBB and eMERGE were combined using the *rma* function from the *metafor* R package under the restricted maximum-likelihood estimator model^37^. We used PheWAS-View to visualize our results^39^. We then created prevalence plots for each significant association. Participants were divided into quintiles based on their PRS, and the percentage of cases for the most significant female health conditions was calculated at each quintile.

### Mendelian Randomization

To identify potential evidence of causality between cardiometabolic phenotypes and female health conditions, we performed one-sample Mendelian randomization (MR) for 29 significant associations from PRS analyses using the *ivreg* function from the *ivpack* R package. Cardiometabolic PRS were used as genetic instruments in the one-sample MR, with birth year and the first five PCs included as covariates. Results were combined through meta-analysis using the *metafor* R package as done in the PRS meta-analysis. Since the low sample size for some of the female health conditions could limit the power of our analyses, we also performed two-sample MR for the same associations using the inverse variance weighted (IVW) method in the *twoSampleMR* package in R^40^. MR sensitivity analyses were conducted using the weighted median and the MR Egger methods through the same package. Genetic instruments for cardiometabolic phenotypes in the two-sample MR were defined as genome-wide significant SNPs in the GWAS summary statistics used to calculate the PRS. SNPs were LD pruned according to LD patterns in the 1000 Genomes HapMap SNPs, and the representative SNPs were included in the analysis. Matching genetic instruments for female health conditions were obtained from publicly available GWAS available through the *twoSampleMR* package (Supplementary Table 2).

### Chronology Analyses

We divided participants into high-risk and low-risk groups according to each cardiometabolic PRS. High PRS was defined as the top quintile (PRS > 80 percentile), and low PRS was defined as the bottom quintile (PRS < 20 percentile). We obtained the age at the first occurrence of each female health condition according to the ICD record. For pregnancy-related conditions, participants were split into three age groups: <25, 25-40, and 40-55. We excluded participants who were over 55 at the first occurrence of pregnancy-related conditions due to low sample sizes and potential errors in diagnosis coding. For all other conditions, participants were split into five different age groups: <25, 25-40, 40-55, 55-70, and >70. We examined the combined case prevalence in PMBB and eMERGE for female health conditions in high and low PRS groups across all age groups.

## Results

### Genetic correlation among cardiometabolic phenotypes and female health conditions

The six cardiometabolic phenotypes were significantly correlated with six different female health conditions for a total of 13 statistically significant correlations (Figure 1A). BMI was significantly correlated with breast cancer (R_g_=-0.179, p=0.011) and PCOS (R_g_=0.4, p=0.007), CAD with breast cancer (R_g_=-0.199, p=0.011), PCOS (R_g_=0.216, p=0.04), and postpartum depression (R_g_=0.204, p=0.025), PP with gestational hypertension (R_g_=0.481, p=0.0025), SBP with breast cancer (R_g_=-0.153, p=0.041) and gestational hypertension (R_g_=0.523, p=0.0017), and T2D with breast cancer (R_g_=-0.196, p=0.0001), excessive fetal growth (R_g_=0.126, p=0.044), gestational diabetes (R_g_=0.529, p=0.011), gestational hypertension (R_g_=0.237, p=0.028), and PCOS (R_g_=0.316, p=0.0093).

**Figure 1:**
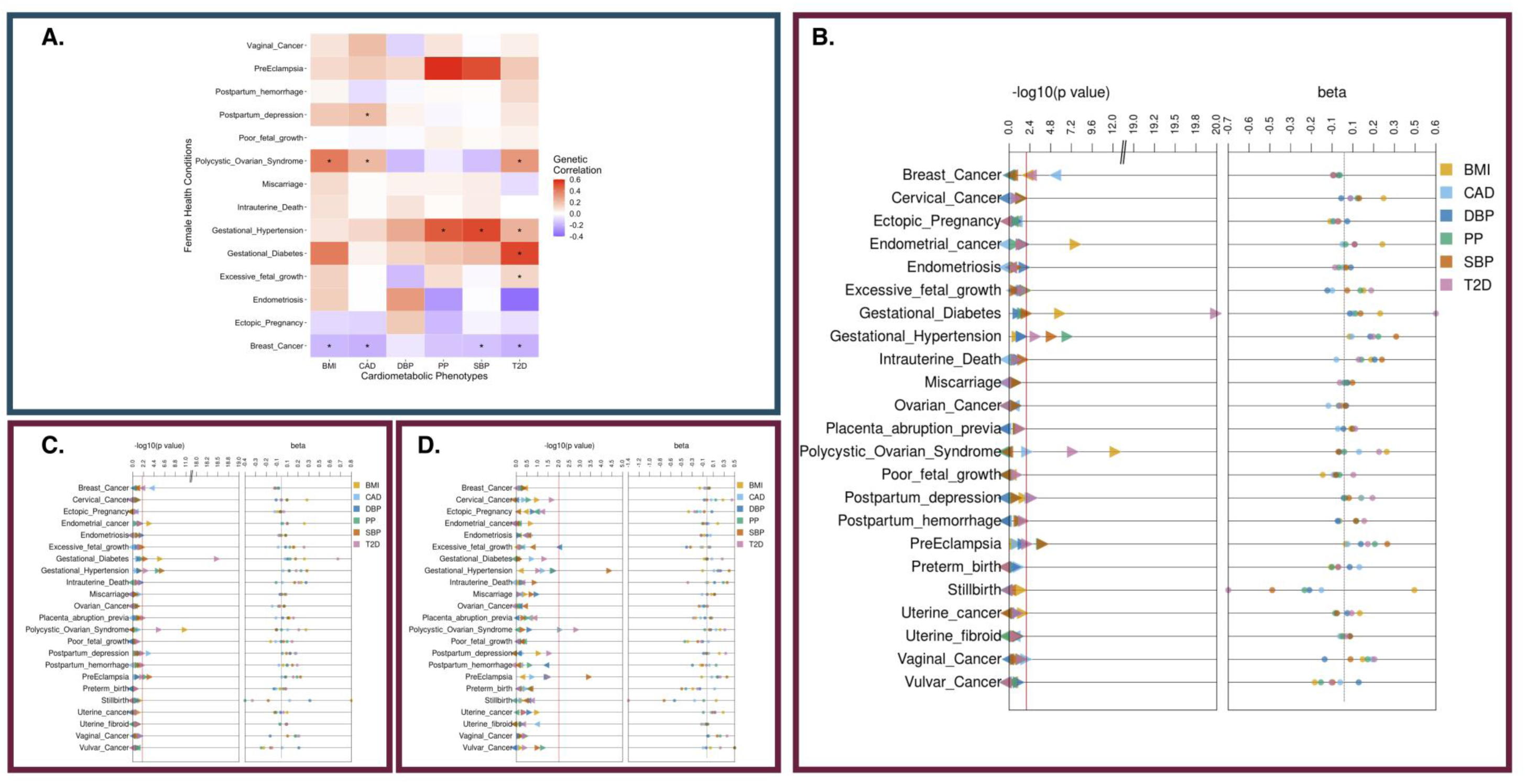
Genetic correlation and the influence of shared genetic burden of cardiometabolic traits and health conditions unique to females. Panel A shows a heatmap of genetic correlation from cross-trait LDSC analyses: Blue represents negative correlation and red represents positive correlation. An * in each box suggests statistical significance of results based on p-values. Panels B, C and D refer to PRS based meta-analyses between cardiometabolic PRS and case/controls status of female-specific diseases in overall, EUR and AFR ancestry individuals respectively. The first panel in these plots corresponds to p-values and second panel represents beta estimates. The color of each point refers to the PRS for cardiometabolic traits.

### PRS performance on primary phenotype

We calculated a PRS for six cardiometabolic phenotypes for individuals in PMBB and eMERGE and checked the distribution of the raw and normalized PRS (Supplementary Figures 1-6). The full model for all PRS generally performed well in predicting the primary phenotype across ancestry groups (Table 3). PRS was significantly (p < 0.05) associated with the primary phenotype for all cardiometabolic phenotypes. The covariate only model (null model) generally performed better in African ancestry individuals than in European individuals, but the PRS only model generally performed better in European ancestry individuals. We calculated the difference between the full and null models for all cardiometabolic PRS in PMBB and eMERGE and found that the PRS improved predictive performance significantly more for European ancestry participants than for African ancestry participants (p = 0.000488, Wilcox signed rank test). As such, the cardiometabolic PRS were more accurate in European ancestry individuals but still significantly improved predictive performance in African ancestry individuals.

**Table 3:** Effect estimates for testing association of PRS with its primary phenotype in eMERGE and PMBB datasets

| Phenotype | Ancestry | N Total | N Cases | R<sup>2</sup> or AUC of Full Model | Beta | SE | P-value | R<sup>2</sup> or AUC of Null Model | R<sup>2</sup> or AUC of PRS only Model |
| --- | --- | --- | --- | --- | --- | --- | --- | --- | --- |
| ***eMERGE*** |  |  |  |  |  |  |  |  |  |
| BMI | All | 39,403 |  | 0.08494 | 2.378 | 0.05044 | 0 | 0.0334 | 0.0593 |
|  | EUR | 31,875 |  | 0.07208 | 2.336 | 0.05144 | 0 | 0.0121 | 0.0583 |
|  | AFR | 5,250 |  | 0.07424 | 2.76 | 0.25 | 5.01E-28 | 0.0529 | 0.0201 |
| CAD | All | 49,171 | 9,597 | 0.784 | 0.3091 | 0.01318 | 1.62E-121 | 0.7745 | 0.5474 |
|  | EUR | 37,003 | 7,524 | 0.7722 | 0.3096 | 0.01427 | 2.82E-104 | 0.7608 | 0.5614 |
|  | AFR | 8,308 | 1,308 | 0.8299 | 0.308 | 0.04789 | 1.24E-10 | 0.827 | 0.5442 |
| DBP (Hypertension) | All | 49,171 | 27,685 | 0.8109 | 0.1387 | 0.01347 | 7.17E-25 | 0.8099 | 0.529 |
|  | EUR | 37,003 | 20,820 | 0.7961 | 0.1464 | 0.01487 | 7.02E-23 | 0.7946 | 0.5339 |
|  | AFR | 8,308 | 4,695 | 0.8634 | 0.1503 | 0.04098 | 0.000245 | 0.8628 | 0.5114 |
| PP (Hypertension) | All | 49,171 | 27,685 | 0.8115 | 0.1644 | 0.01344 | 2.15E-34 | 0.8099 | 0.526 |
|  | EUR | 37,003 | 20,820 | 0.7969 | 0.1712 | 0.01438 | 1.13E-32 | 0.7946 | 0.5273 |
|  | AFR | 8,308 | 4,695 | 0.8629 | 0.09689 | 0.04871 | 0.0467 | 0.8628 | 0.5049 |
| SBP (Hypertension) | All | 49,171 | 27,685 | 0.8131 | 0.2977 | 0.01665 | 1.86E-71 | 0.8099 | 0.5352 |
|  | EUR | 37,003 | 20,820 | 0.7992 | 0.3156 | 0.01793 | 2.28E-69 | 0.7946 | 0.5428 |
|  | AFR | 8,308 | 4,695 | 0.8637 | 0.2845 | 0.06227 | 4.91E-06 | 0.8628 | 0.5116 |
| T2D | All | 49,171 | 12,403 | 0.7193 | 0.6334 | 0.01914 | 3.02E-240 | 0.691 | 0.6163 |
|  | EUR | 37,003 | 8,311 | 0.6869 | 0.6311 | 0.0205 | 3.56E-208 | 0.6414 | 0.6033 |
|  | AFR | 8,308 | 2,725 | 0.7795 | 0.6318 | 0.06546 | 4.83E-22 | 0.7728 | 0.5597 |
| ***PMBB*** |  |  |  |  |  |  |  |  |  |
| BMI | All | 20,209 |  | 0.1542 | 2.658 | 0.08244 | 2.05E-222 | 0.1108 | 0.1417 |
|  | EUR | 12,344 |  | 0.07473 | 2.621 | 0.08679 | 2.66E-193 | 0.006422 | 0.06782 |
|  | AFR | 6,744 |  | 0.03203 | 2.708 | 0.2089 | 5.75E-38 | 0.008037 | 0.02711 |
| CAD | All | 21,837 | 3,002 | 0.8235 | 0.3571 | 0.02354 | 5.73E-52 | 0.8148 | 0.5619 |
|  | EUR | 13,515 | 1,956 | 0.8303 | 0.3894 | 0.02732 | 4.44E-46 | 0.8185 | 0.5929 |
|  | AFR | 7,039 | 955 | 0.799 | 0.2566 | 0.05214 | 8.60E-07 | 0.7955 | 0.5389 |
| DBP | All | 21,612 |  | 0.0425 | 1.036 | 0.06661 | 3.05E-54 | 0.03183 | 0.02905 |
|  | EUR | 13,343 |  | 0.01534 | 0.9964 | 0.08064 | 7.01E-35 | 0.004144 | 0.01213 |
|  | AFR | 6,994 |  | 0.04559 | 1.104 | 0.129 | 1.35E-17 | 0.03571 | 0.01375 |
| PP (Hypertension) | All | 21,837 | 10,278 | 0.8125 | 0.1942 | 0.02328 | 7.18E-17 | 0.8112 | 0.602 |
|  | EUR | 13,515 | 5,640 | 0.7797 | 0.1918 | 0.02638 | 3.53E-13 | 0.7778 | 0.5272 |
|  | AFR | 7,039 | 4,286 | 0.8389 | 0.1886 | 0.05376 | 0.00045 | 0.8383 | 0.5391 |
| SBP (Hypertension) | All | 21,837 | 10,278 | 0.8144 | 0.327 | 0.02523 | 2.08E-38 | 0.8112 | 0.6097 |
|  | EUR | 13,515 | 5,640 | 0.7813 | 0.279 | 0.02846 | 1.11E-22 | 0.7778 | 0.5397 |
|  | AFR | 7,039 | 4,286 | 0.8415 | 0.4505 | 0.05923 | 2.81E-14 | 0.8383 | 0.5649 |
| T2D | All | 21,837 | 4,388 | 0.754 | 0.8136 | 0.03523 | 5.49E-118 | 0.7287 | 0.6713 |
|  | EUR | 13,515 | 1,969 | 0.712 | 0.8418 | 0.04533 | 5.55E-77 | 0.6634 | 0.6323 |
|  | AFR | 7,039 | 2,221 | 0.7342 | 0.7896 | 0.05947 | 3.11E-40 | 0.7168 | 0.5919 |

\*\*BMI= Body Mass Index; CAD= Coronar artery disease, T2D=Type 2 Diabetes; DBP= Diastolic Blood Pressure; SBP= Systolic Blood Pressure; PP= Pulse Pressure; EUR= European ancestry; AFR= African ancestry; Beta coefficients and standard errors are per SD PRS in the full model. Covariates included: birth year and the first five PCs.

### Association of cardiometabolic PRS association and female health conditions

We detected numerous associations between cardiometabolic PRS and female health conditions in the meta-analyzed results (Figure 1B-D). 29 associations were statistically significant after correcting for multiple hypothesis burden through FDR significance (Table 4). Most of these associations were also significantly associated in both PMBB and eMERGE separately (Supplementary Figures 7-12). In the meta-analysis, for all and European ancestry individuals, the PRS_BMI_ was significantly associated with endometrial cancer (beta_all_=0.24, se_all_=0.046, p_all_=9.4×10^-8^; beta_eur_=0.28, se_eur_=0.087, p_eur_=0.0015), gestational diabetes (beta_all_=0.23, se_all_=0.051, p_all_ =6×10^-6^; beta_eur_=0.28, se_eur_=0.062, p_eur_=8.7×10^-6^), and PCOS ((beta_all_=0.27, se_all_=0.039, p_all_=2.4×10^-12^; beta_eur_=0.29, se_eur_=0.045 p_eur_=6.8×10^-11^). The PRS_BMI_ was also significantly associated with breast cancer in all individuals (beta_all_=-0.071, se_all_=0.022, p_all_=0.0016). These results suggest that an increased genetic burden for obesity and high BMI also increases the risk for endometrial cancer, gestational diabetes, and PCOS but decreases the risk for breast cancer. The PRS_CAD_ was also significantly associated with breast cancer for all and European ancestry individuals (beta_all_=-0.072, se_all_=0.015, p_all_=1×10^- 6^; beta_eur_=-0.07, se_all_=0.017, p_eur_=3.1×10^-5^). This highly significant negative association suggests that individuals, particularly those of European ancestry, with high PRS_CAD_ are at relatively lower risk for breast cancer compared to individuals with low PRS_CAD_. The PRS_T2D_ was also significantly associated with gestational diabetes in all and European ancestry individuals (beta_all_=0.58, se_all_=0.063, p_all_=1.2×10^-20^; beta_eur_=0.68, se_eur_=0.076, p_eur_=3.9×10^-19^) and with PCOS in all, European ancestry, and African ancestry individuals (beta_all_=0.22, se_all_=0.043, p_all_=1.9×10^-7^; beta_eur_=0.21, se_eur_=0.049, p_eur_=1.6×10^-5^; beta_afr_=0.32, se_afr_=0.1, p_afr_=0.0021). The link between T2D and these female related phenotypes is well known, and our results support a potential genetic basis linking these phenotypes^18, 19^. Among all and European ancestry individuals, the PRS_T2D_ was also significantly associated with gestational hypertension (beta_all_=0.18, se_all_=0.064, p_all_=0.0041; beta_eur_=0.18, se_eur_=0.067, p_eur_=0.008) and breast cancer (beta_all_=-0.073, se_all_=0.021, p_all_=0.00066; beta_eur_=-0.079, se_eur_=0.026, p_eur_=0.0027).

**Table 4:**
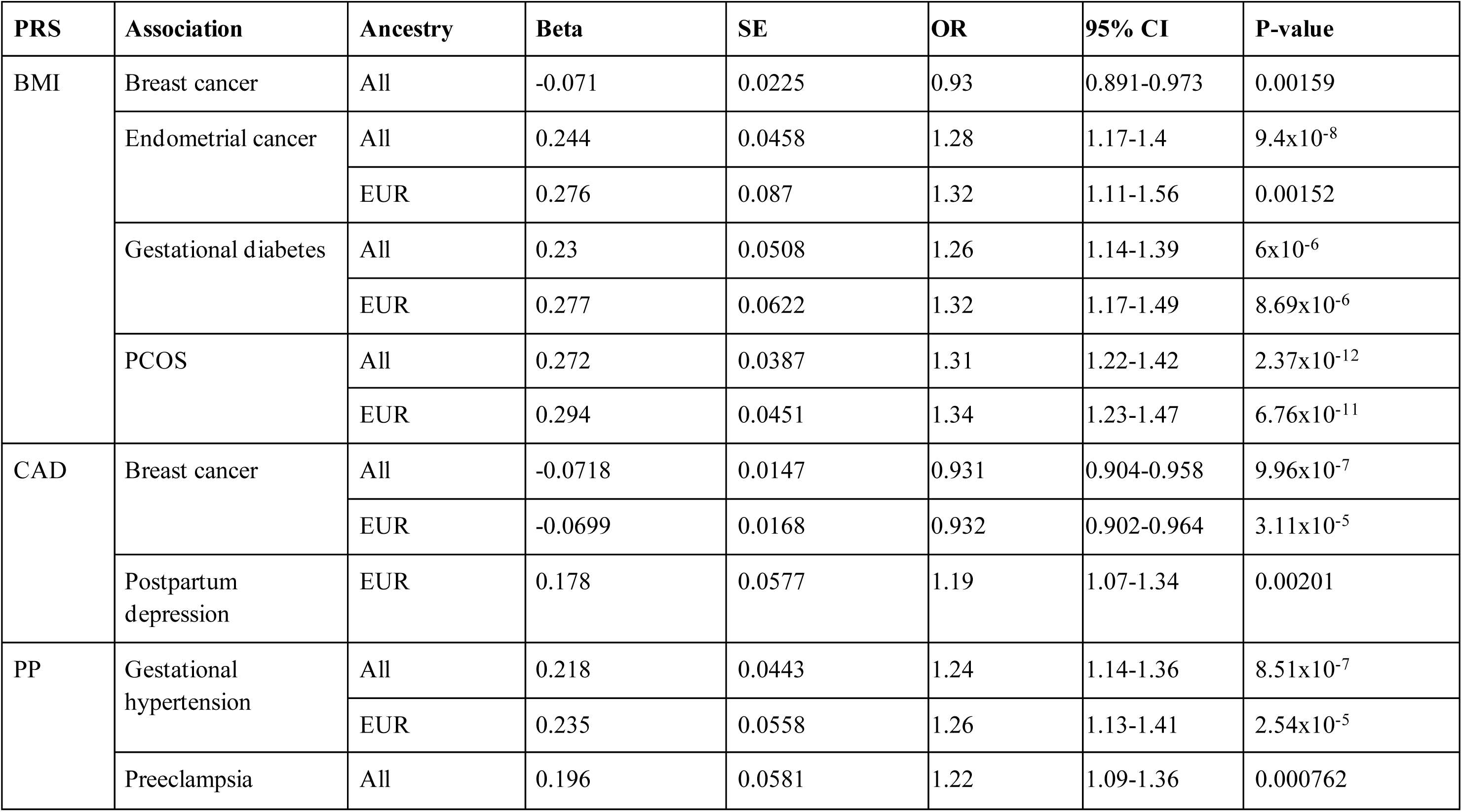
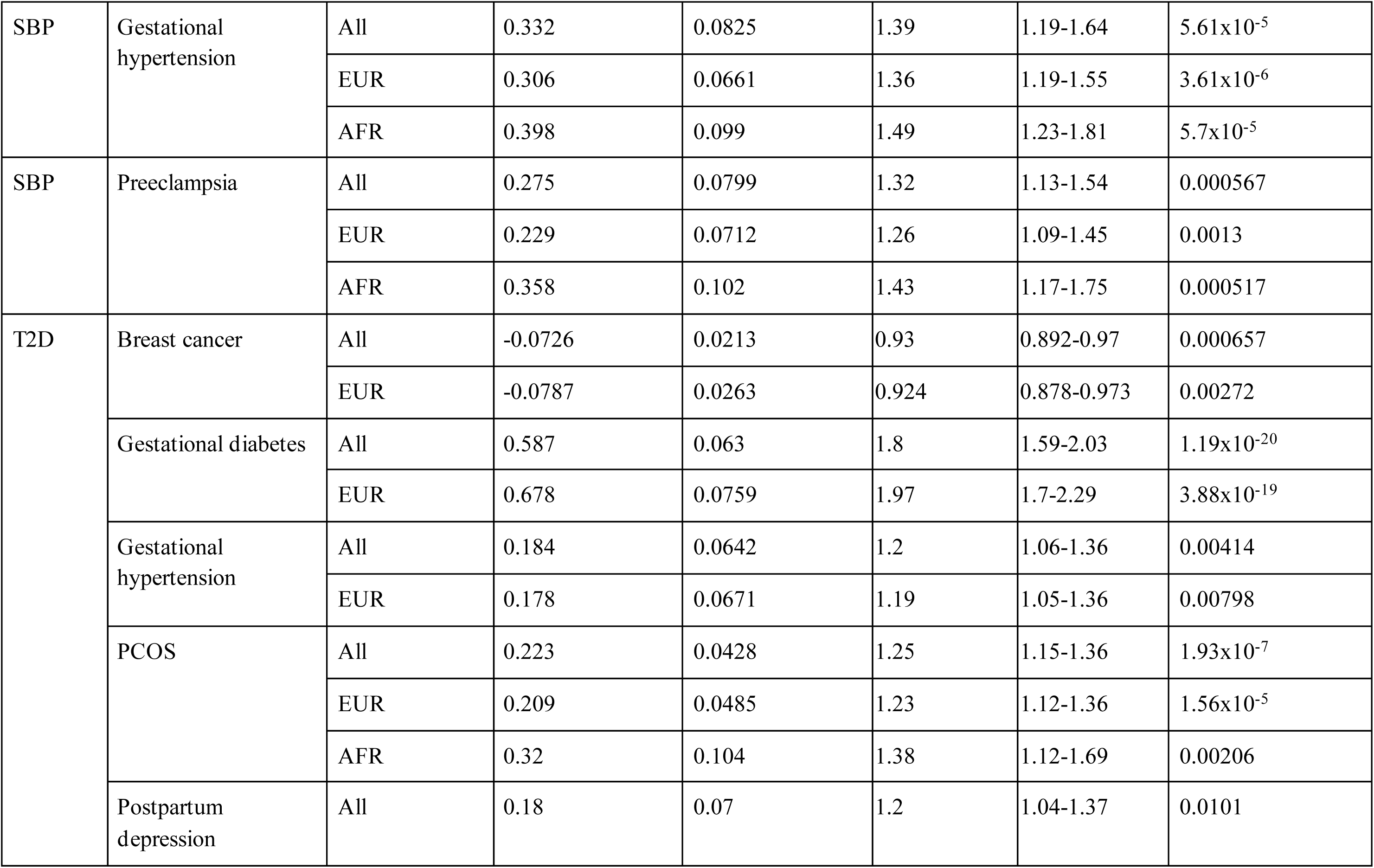
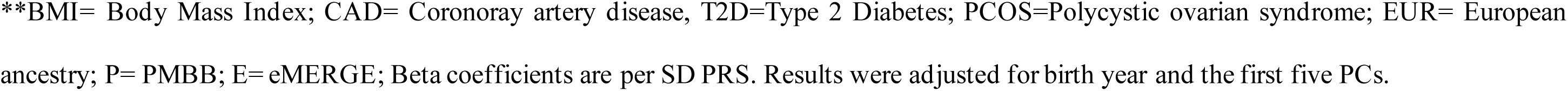
FDR-significant (FDR P-value < 0.05) associations between cardiometabolic PRS and female health conditions identified in the PMBB and eMERGE meta-analysis

| PRS | Association | Ancestry | Beta | SE | OR | 95% CI | P-value |
| --- | --- | --- | --- | --- | --- | --- | --- |
| BMI | Breast cancer | All | -0.071 | 0.0225 | 0.93 | 0.891-0.973 | 0.00159 |
|  | Endometrial cancer | All | 0.244 | 0.0458 | 1.28 | 1.17-1.4 | 9.4x10<sup>-8</sup> |
|  |  | EUR | 0.276 | 0.087 | 1.32 | 1.11-1.56 | 0.00152 |
|  | Gestational diabetes | All | 0.23 | 0.0508 | 1.26 | 1.14-1.39 | 6x10<sup>-6</sup> |
|  |  | EUR | 0.277 | 0.0622 | 1.32 | 1.17-1.49 | 8.69x10<sup>-6</sup> |
|  | PCOS | All | 0.272 | 0.0387 | 1.31 | 1.22-1.42 | 2.37x10<sup>-12</sup> |
|  |  | EUR | 0.294 | 0.0451 | 1.34 | 1.23-1.47 | 6.76x10<sup>-11</sup> |
| CAD | Breast cancer | All | -0.0718 | 0.0147 | 0.931 | 0.904-0.958 | 9.96x10<sup>-7</sup> |
|  |  | EUR | -0.0699 | 0.0168 | 0.932 | 0.902-0.964 | 3.11x10<sup>-5</sup> |
|  | Postpartum depression | EUR | 0.178 | 0.0577 | 1.19 | 1.07-1.34 | 0.00201 |
| PP | Gestational hypertension | All | 0.218 | 0.0443 | 1.24 | 1.14-1.36 | 8.51x10<sup>-7</sup> |
|  |  | EUR | 0.235 | 0.0558 | 1.26 | 1.13-1.41 | 2.54x10<sup>-5</sup> |
|  | Preeclampsia | All | 0.196 | 0.0581 | 1.22 | 1.09-1.36 | 0.000762 |
| SBP | Gestational hypertension | All | 0.332 | 0.0825 | 1.39 | 1.19-1.64 | 5.61x10<sup>-5</sup> |
|  |  | EUR | 0.306 | 0.0661 | 1.36 | 1.19-1.55 | 3.61x10<sup>-6</sup> |
|  |  | AFR | 0.398 | 0.099 | 1.49 | 1.23-1.81 | 5.7x10<sup>-5</sup> |
| SBP | Preeclampsia | All | 0.275 | 0.0799 | 1.32 | 1.13-1.54 | 0.000567 |
|  |  | EUR | 0.229 | 0.0712 | 1.26 | 1.09-1.45 | 0.0013 |
|  |  | AFR | 0.358 | 0.102 | 1.43 | 1.17-1.75 | 0.000517 |
| T2D | Breast cancer | All | -0.0726 | 0.0213 | 0.93 | 0.892-0.97 | 0.000657 |
|  |  | EUR | -0.0787 | 0.0263 | 0.924 | 0.878-0.973 | 0.00272 |
|  | Gestational diabetes | All | 0.587 | 0.063 | 1.8 | 1.59-2.03 | 1.19x10<sup>-20</sup> |
|  |  | EUR | 0.678 | 0.0759 | 1.97 | 1.7-2.29 | 3.88x10<sup>-19</sup> |
|  | Gestational hypertension | All | 0.184 | 0.0642 | 1.2 | 1.06-1.36 | 0.00414 |
|  |  | EUR | 0.178 | 0.0671 | 1.19 | 1.05-1.36 | 0.00798 |
|  | PCOS | All | 0.223 | 0.0428 | 1.25 | 1.15-1.36 | 1.93x10<sup>-7</sup> |
|  |  | EUR | 0.209 | 0.0485 | 1.23 | 1.12-1.36 | 1.56x10<sup>-5</sup> |
|  |  | AFR | 0.32 | 0.104 | 1.38 | 1.12-1.69 | 0.00206 |
|  | Postpartum depression | All | 0.18 | 0.07 | 1.2 | 1.04-1.37 | 0.0101 |

\*\*BMI= Body Mass Index; CAD= Coronaray artery disease, T2D=Type 2 Diabetes; PCOS=Polycystic ovarian syndrome; EUR= European ancestry; P= PMBB; E= eMERGE; Beta coefficients are per SD PRS. Results were adjusted for birth year and the first five PCs.

The three-blood pressure traits PRS (PRS_DBP_, PRS_SBP_, and PRS_PP_) showed varying significant associations with gestational hypertension and preeclampsia in the PMBB and eMERGE meta-analysis. There was no sign of association between the PRS_DBP_ and these phenotypes in the meta-analysis. However, the PRS_DBP_ was significantly associated with gestational hypertension in PMBB all and African ancestry individuals (p=1×10^-05^, 0.00041) and nominally associated (p<0.01) with preeclampsia in African ancestry participants (p=0.0063). The PRS_PP_ was significantly associated with gestational hypertension for all and European ancestry participants (beta_all_=0.22, se_all_=0.044, p_all_=8.5×10^-7^; beta_eur_=0.24, se_eur_=0.056, p_eur_=2.5×10^-5^) and with preeclampsia for all participants (beta_all_=0.2, se_all_=0.058, p_all_=0.00076). The PRS_SBP_ was significantly associated with gestational hypertension and preeclampsia in all, European ancestry, and African ancestry individuals (Gestational hypertension: beta_all_=0.33, se_all_=0.083, p_all_=5.6×10^-5^; beta_eur_=0.31, se_all_=0.066, p_eur_=3.6×10^-6^; beta_afr_=0.4, se_afr_=0.099, p_afr_=5.7×10^-5^; Preeclampsia: beta_all_=0.28, se_all_=0.08, p_all_=0.00057; beta_eur_=0.23, se_eur_=0.071, p_eur_=0.0013; beta_afr_=0.36, se_afr_=0.1, p_afr_=0.00052).

For each FDR-significant association in both PMBB and eMERGE, we looked at the case prevalence of the female health condition per the associated PRS quintile (Figure 2B and Supplementary Figures 13-17). The trends matched the results we obtained from the association analyses across ancestry groups and in both datasets. Distribution of the number of cases increased as the PRS percentile increased for the positive associations in the association analysis, such as PCOS with the PRS_BMI_. The number of cases for breast cancer decreased when increasing the PRS_CAD_ percentile, matching the inverse association between breast cancer and the PRS_CAD_ (Figure 2B).

**Figure 2:**
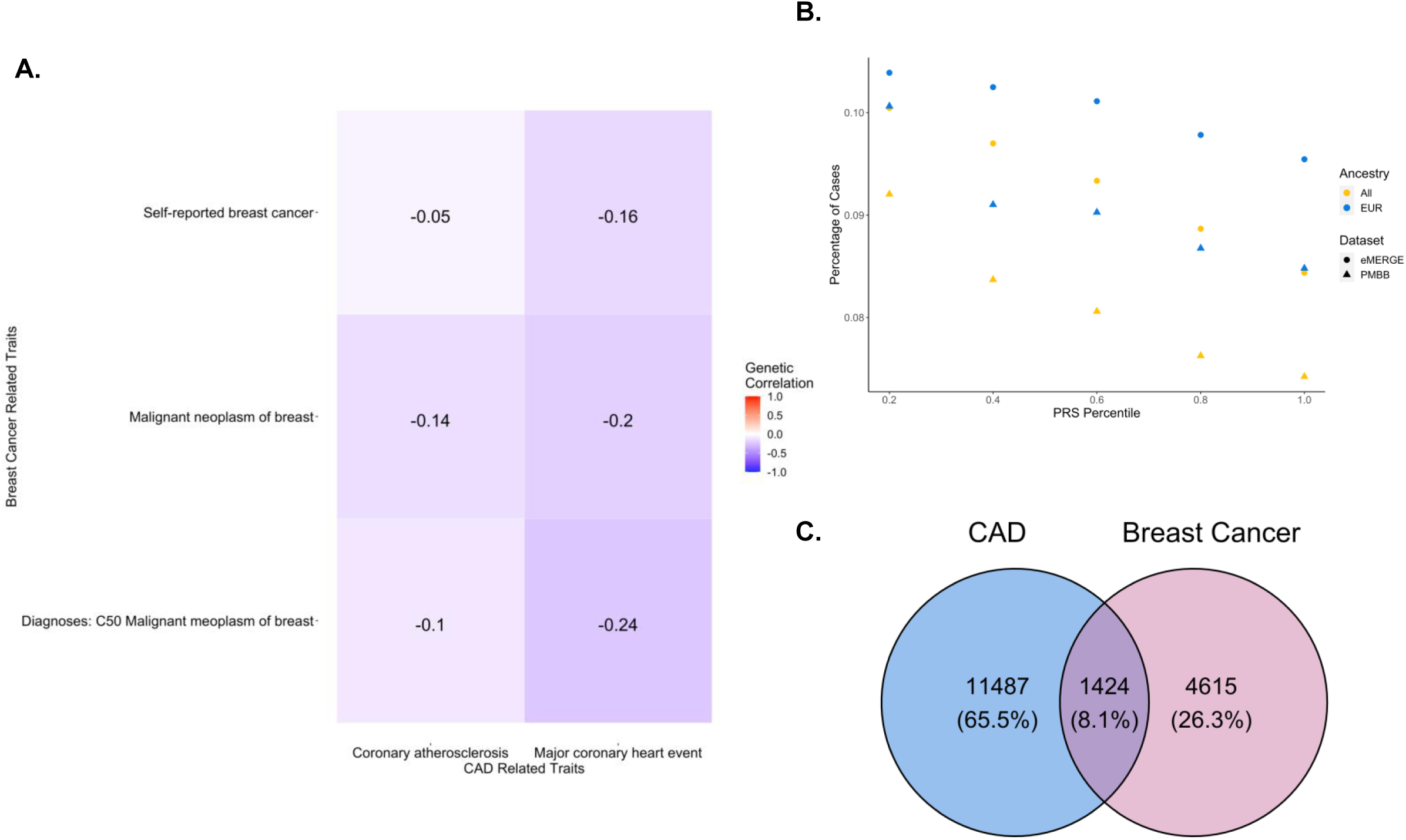
Inverse relationship between Coronary Artery Disease (CAD) and Breast Cancer. Panel A shows the heatmap of negative genetic correlation between CAD and breast cancer from the UKBB Genetic Correlation Browser dataset. The gradient of color refers to positive (red) to negative (blue) correlation, and the text in each box refers to the genetic correlation coefficient. Panel B shows the distribution of breast cancer per each PRS_CAD_ quintile. The x-axis represents each PRS quintile, and the y-axis is the disease prevalence. The color of each point refers to the ancestry group (All, European, and African), and the shape indicates the target dataset (eMERGE (circle) or PMBB (triangle)). Panel C is a Venn diagram representing the overlap of CAD and breast cancer cases in PMBB and eMERGE datasets based on disease diagnosis.

### Association of PRS_CAD_ and breast cancer

To validate the inverse association between PRS_CAD_ and breast cancer, we examined the genetic correlation between CAD and breast cancer from the UKBB Genetic Correlation Browser (Figure 2A). The genetic correlation between I9_CHD (Major coronary heart disease event) and C50 (Diagnoses-main ICD10: C50 Malignant neoplasm of breast) was significantly negative (R_g_=-0.24, p =0.0325).

Additionally, to evaluate the risk of ascertainment biases and the reporting of comorbidities in the EHR, we preformed the associations of CAD PRS with breast cancer in females who are not diagnosed with CAD (n=61,201). In the individuals who were not diagnosed with CAD, we observed slightly less significant association between breast cancer and CAD (beta_all_=-0.0484, p_all_=0.0045; beta_eur_=-0.0464, p_eur_=0.0112) than in the full set of individuals. These associations attenuated but were not completely diminished when evaluating participants who were not diagnosed with CAD. Next, given the longitudinal nature of availability of comorbidities in the EHR, we aimed to measure the impact of risk of CAD on the first reported incidence of CAD and breast cancer in females. The overlap on individuals who were diagnosed with CAD and breast cancer in our study population suggests that females with a diagnosis of CAD are less likely to be diagnosed or report a history of breast cancer (Figure 2C).

### Mendelian Randomization

We performed one-sample MR in PMBB and eMERGE for the 29 associations that were FDR-significant and combined them through a meta-analysis (Figure 3A). The majority of analyses were significant (p < 0.05), and the direction of the estimated beta coefficient for each association aligned with the direction of effect seen in the PRS association analysis. Our results support a potential causal relationship between many cardiometabolic phenotypes and female health conditions, with the most significant associations being between T2D and gestational diabetes (All: p=1.5×10^-11^, beta=1.52, se=0.23; EUR: p=2.4×10^-10^, beta=1.78, se=0.28) and BMI and PCOS (All: p=1.1×10^-8^, beta=0.0021, se=0.00037; EUR: p=3.6×10^-8^, beta=0.0021, se=0.0038).

**Figure 3:**
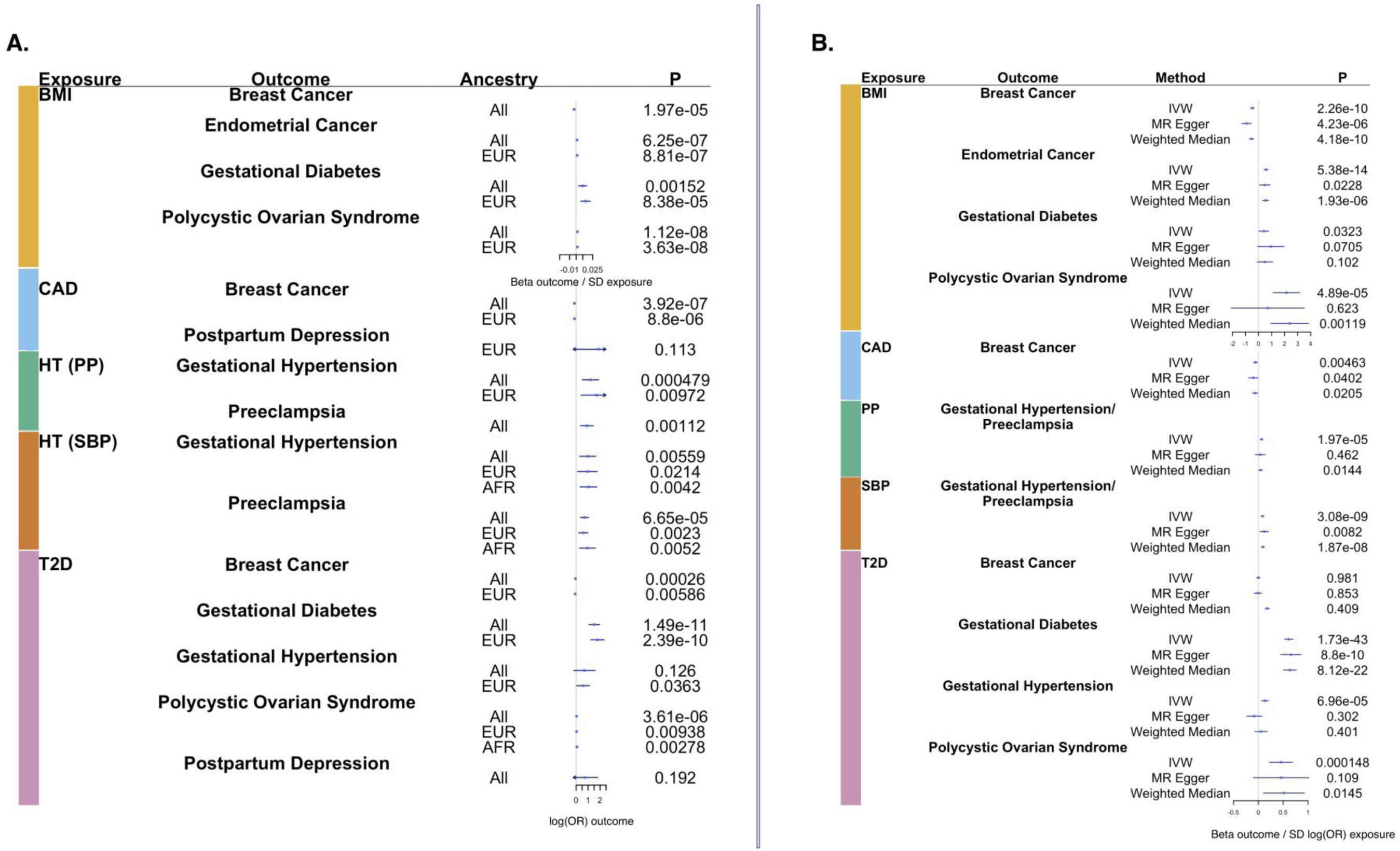
Mendelian Randomization (MR) results for 29 significant PRS based associations. This figure shows forest plots of association between female health conditions as outcomes and cardiometabolic phenotypes as exposures in a one-sample MR analyses shown in Panel A and two-sample MR analyses shown in Panel B. Genetic instruments are the cardiometabolic PRS in the one-sample MR and genome-wide significant SNPs from GWAS in the two-sample MR. In panel A, each point refers to beta outcome/SD exposure for PRS_BMI_ and log(OR) outcome for all other exposure variables for each tests performed as separated by ancestry. In panel B, each point refers to beta outcome/SD exposure for BMI and beta outcome/SD log(OR) exposure for all other variables across all methods used in sensitivity analyses for two-sample MR. P-values are reported in last column in both panels.

We also performed two-sample MR to leverage the power of larger GWAS for our female health conditions (Figure 3B). Most analyses were significant when using the IVW method but became less significant when using the MR Egger and the weighted median methods. Some associations remained significant for all three methods, particularly CAD with breast cancer (IVW: beta=-0.0587, se=0.021, p=0.00463; MR Egger: beta=-0.102, se=0.049, p=0.04; Weighted median: beta=-0.0642, se=0.028, p=0.021) and T2D and gestational diabetes (IVW: beta=0.613, se=0.044, p=1.73×10^-43^; MR Egger: beta=0.656, se=0.11, p=8.8×10^-10^; Weighted median: beta=0.635, se=0.066, p=8.12×10^-22^).

### Investigation of the role of population stratification

Population stratification could have confounded our results in the PRS association and the MR analyses. Therefore, we tested the association of the PRS with the PCs (Supplementary Table 3). The high R^2^ for most cardiometabolic PRS shows that a large amount of the variance in the PRS could be explained by the PCs when considering all multi-ancestry variables, and the R^2^ decreases when analyzing only European or African ancestry groups. The PCs explained more of the variance in PRS for African ancestry individuals than European ancestry individuals, a finding that suggests there is more population stratification among our African ancestry individuals and that there could be confounding factors influencing some results. To overcome these biases, we accounted for PCs in both our PRS association analyses and the one-sample MR analyses. Additionally, we ran one-sample MR without adjusting for the covariates (PCs and birth year) and compared the results with the adjusted ones (Supplementary Table 4).

### Chronology analyses

EHR is a powerful resource to visualize the landscape of events in a patient ’s diagnosis. Continuing on the theme of genetic correlation and the influence of shared genetic burden of cross-trait analyses, we investigated if an individual’s genetic burden for cardiometabolic diseases may also affect the time at which they develop certain female health conditions. Case prevalence for female health conditions was generally higher in the high PRS group in the younger age groups (Figure 4 and Supplementary Figures 18-22). Many pregnancy-related phenotypes were most prevalent in the 25-40 years age group, but for participants who developed those phenotypes in the <25 years age group, the relative difference in prevalence between the high-risk and low-risk PRS group was larger. The cardiometabolic PRS used to define high-risk and low-risk groups did not seem to affect case prevalence for most phenotypes, but phenotypes found to be associated with specific cardiometabolic PRS in the association analysis did show differences across PRS. Gestational hypertension and preeclampsia were generally more prevalent in the younger age groups (<25 and 25-40) for participants with high blood pressure PRS than for participants with high PRS for other phenotypes. Similar trends were observed for other traits, such as the PRS_T2D_ with gestational diabetes and the PRS_BMI_ with PCOS. High prevalence of endometriosis and uterine fibroid cases were also observed in individuals with high-risk PRS, particularly in the age group 25-40.

**Figure 4:**
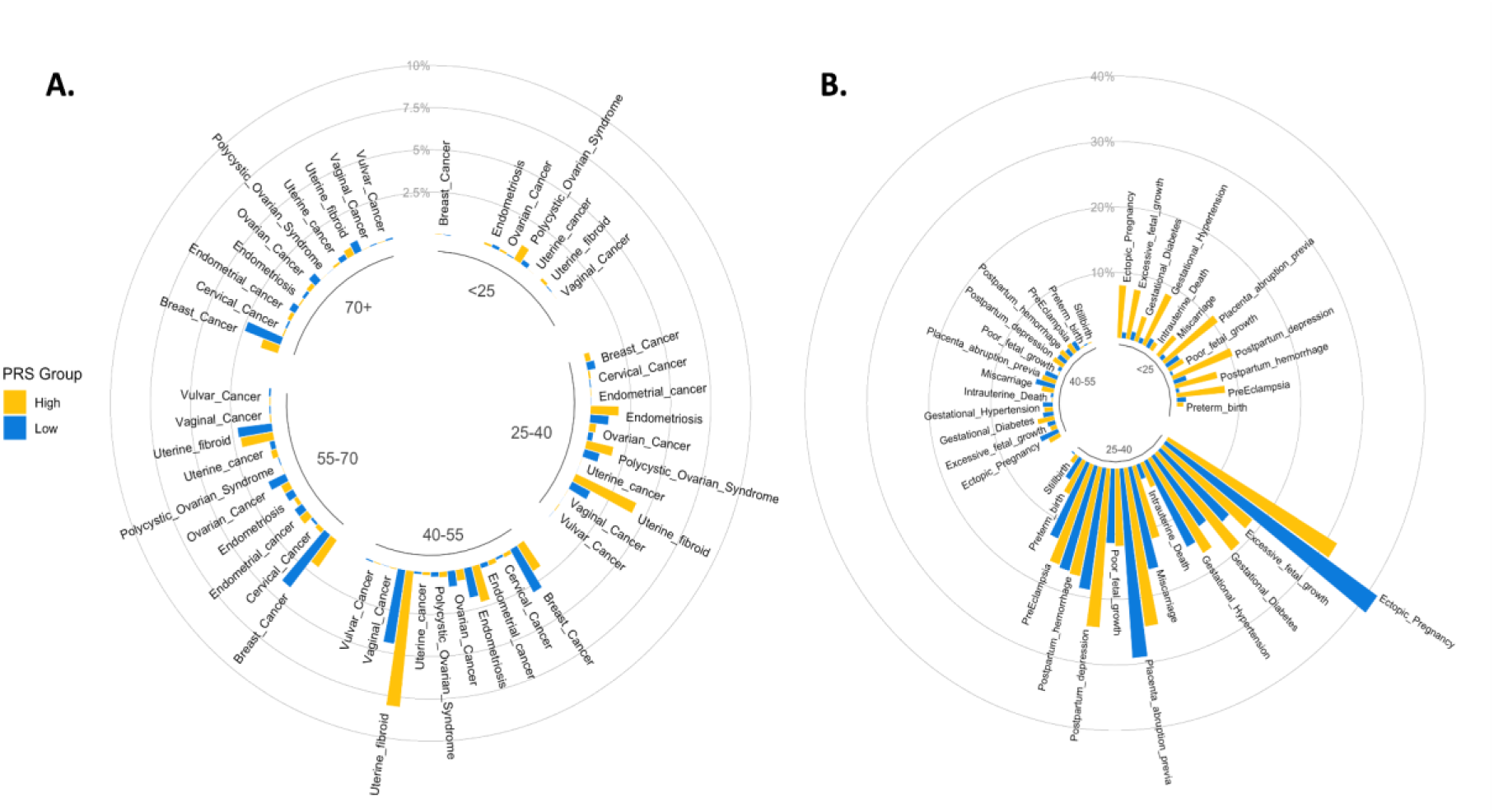
Chronology analyses for the visualization of events from the EHR. Circular plot showing disease prevalence among high PRS_BMI_ (in yellow) and low PRS_BMI_ (in blue) categories. General female health conditions are shown in panel A and pregnancy and childbirth-related phenotypes are shown in panel B. The circular plots are divided into five age categories (<25, 25-40, 40-55, 55-70, and >70) for general female health conditions and three age categories (<25, 25-40, and 40-55) for pregnancy-related phenotypes

## Discussion

We have demonstrated high genetic correlation and shared genetic burden between cardiometabolic traits and female health conditions spanning across many obstetrics and gynecological disorders. Additionally, this study shows that genome-wide PRS of cardiometabolic traits has potential translational effects in health conditions unique to females. Initially, we calculated the genetic correlation between phenotypes related to cardiometabolic diseases and traits with female health conditions. Genetic correlation analyses suggested a high overlap among the shared genetic etiology of all phenotypes tested in this study. We further investigated the effect of this shared genetic burden by generating PRS for cardiometabolic phenotypes and testing their association to various female-specific disease conditions. PRS was generally predictive of the primary phenotype, and we identified several FDR-significant associations with female health conditions that were statistically significant after meta-analyzing both datasets. Prior research has established relationships (mainly non-genetic factors) between many of these associations, such as BMI with PCOS and T2D with gestational diabetes^17, 18^. Epidemiological studies have also shown evidence that people with obesity are at high risk for endometrial cancer^41^. The associations identified in this study between female health conditions and cardiometabolic PRS suggest genetic basis for these cross-trait categories. In addition, we performed Mendelian randomization and identified potential causal relationships.

Unexpectedly, all of our analyses showed that the PRS_CAD_ was inversely associated with breast cancer in European ancestry participants. CAD and breast cancer share many common risk factors, such as smoking and diet^42^. However, our results suggest that a high genetic burden for CAD is protective for breast cancer. Participants in our cohorts with high PRS_CAD_ had lower incidence of breast cancer. When we evaluated the individuals with high PRS_CAD_ but no CAD diagnoses, we still saw a moderate but decreased risk for breast cancer. Several factors might contribute to these associations. First, these patterns might reflect ascertainment biases and competing risks. Individuals with higher risk of CAD likely live shorter lives and thus are not diagnosed with breast cancer. Second, the treatments and drugs given for treating CAD might also protect against breast cancer. Lastly, genetic mechanisms predisposing individuals to CAD might show protective effects for breast cancer. Briefly, among the SNPs that passed genome-wide significance in the CAD GWAS we used to calculate the PRS_CAD_, 125/220 SNPs had opposite directions of effects in the breast cancer GWAS we used in the two-sample MR. For example, *rs1011970* (CAD: beta=-0.0407, p=6.94×10^-10^; breast cancer: beta=0.053, p=3.1×10^-5^) is a known risk variant for both CAD and breast cancer and maps to the *CDKN2B-AS1* gene, which helps silence genes epigenetically in the *CDKN2A-CDKN2B* cluster^43^. *CDKN2B-AS1* knockdown suppresses breast cancer progression, and decreased expression of *CDKN2B* increases development of atherosclerotic plaques^44–46^. Variants that promote decreased *CDKN2B-AS1* expression could increase breast cancer risk but increase expression of *CDKN2B* and subsequently decrease CAD risk. A recent study also shows this protective effect of a PRS_CAD_ on breast cancer, and more research should be undertaken to uncover the mechanisms behind this association^47^.

The differences between the three PRS_blood pressure_ suggests that analyzing the genetic burden for all three blood pressure traits might elucidate the understanding of hypertension during pregnancy. Only the PRS_SBP_ was significantly associated with gestational hypertension and preeclampsia in African ancestry individuals. Prior studies have identified variability in the predictive performance when using different blood pressure measurements to predict hypertension and other diseases, and our results warrant further studies to replicate this association in other large multi-ancestry datasets to reach a more definite conclusion on the role blood pressure measurements play in predicting hypertensive diseases during pregnancy^48, 49^.

To further investigate the effect of cardiometabolic genetic burden on conditions unique to females, we drew information from EHRs to capture the landscape of trajectories of diseases presented at different ages. We found that a high genetic burden for most cardiometabolic phenotypes could increase risk of developing female health conditions at earlier ages, even if there was no overall association between the PRS and the condition. For example, the PRS_BMI_ was not associated with many pregnancy-related complications such as gestational hypertension and ectopic pregnancy. However, in the youngest age group (<25), these phenotypes were more prevalent in the high PRS_BMI_ group than the low PRS_BMI_ group. Similarly, there was no association overall between the PRS_SBP_ and PCOS or endometriosis, but participants who developed PCOS before 25 years old or endometriosis between 25-40 years old were more likely to have high PRS. The difference in case prevalence between high and low PRS groups for these phenotypes decreased in older age groups. Patients with high genetic burden for cardiometabolic phenotypes could be at higher risk to develop female health conditions at younger ages. These findings can be particularly of importance to prioritize patients for early screening.

The limitations of our study point to many promising future directions. First, our study establishes a link between the genetic burden for different cardiometabolic phenotypes and several diseases unique to females. However, we estimate the genetic burden by using PRS alone, which are calculated based on common variants and do not include the effects of other genetic risk factors such as rare variants and copy number variations. Second, our analysis does not consider clinical or environmental factors that could influence the associations between cardiometabolic and female health conditions. We found that population stratification is potentially present in our datasets and accounted PCs in our analyses accordingly. However, important social and environmental risk factors such as education level and socioeconomic status were missing and could not be properly accounted for in our current analyses. Our focus on PRS alone reflects the current limitations of methods for multi-modal risk model predictions. Current widespread interest in building integrative risk models suggests that this gap can be closed in the near future^27, 50^. Furthermore, the weaker performance of PRS in African ancestry individuals contributed to the lack of power to identify African ancestry specific associations and suggests the urgent need for expanding studies to include more racially and ethnically diverse cohorts.

The low sample size for some conditions may have limited the power of our one-sample MR analyses. In addition, while our two-sample MR uses GWAS from larger cohorts, they were performed on populations of primarily European ancestry and thus lack the power to detect causality for the non-European ancestry participants in our cohort. The effects we see in our MR results may be biased due to horizontal pleiotropy, in which genetic variants associated with the exposure (cardiometabolic phenotypes) affect other traits that subsequently influence the outcome (female health conditions). The effects of pleiotropy can be seen from the decreased statistical significance when using the weighted median and MR Egger methods in the two-sample MR. For example, the relationship between T2D and PCOS became insignificant when using the MR Egger method (p = 0.109) compared to the IVW method (p = 0.000148). These methods account for pleiotropy and other confounding factors but may not have captured all their effects.

EHRs are particularly advantageous in investigating disease trajectories and progression. Our analyses in this study provided a big picture visualization of the burden of early diagnoses of disease unique to females at early ages among the high cardiometabolic PRS group. However, these analyses might point to the ascertainment biases of EHR and including confounding factors for statistically informed findings. This study illustrates the influence of cardiometabolic genetic burden on diverse phenotypes. Our findings serve as the initial basis for presenting the clinical utility of cardiometabolic PRS as non-modifiable risk factors for screening and early diagnostic tools for a variety of obstetric and gynecological conditions. To improve the power of cardiometabolic PRS for predicting risk for female health conditions and better understand the relationship between these phenotypes, future studies should incorporate PRS with other genetic and non-genetic risk factors and study their effects on larger and more diverse multi-ancestry populations.

## Data Availability

All data produced in this study are available upon  request to the authors. Summary level data will also be available upon request.

## Acknowledgements

We acknowledge the Penn Medicine BioBank (PMBB) for providing data and thank the patient-participants of Penn Medicine who consented to participate in this research program. We would also like to thank the Penn Medicine BioBank team and Regeneron Genetics Center for providing genetic variant data for analysis. The PMBB is approved under IRB protocol #813913 and supported by Perelman School of Medicine at University of Pennsylvania. Approval for PMBB was given from the University of Pennsylvania Institutional Review Board. The PMBB study has been determined to pose minimal risk to subjects.

## Sources of Funding

The eMERGE Network was initiated and funded by NHGRI through the following grants:

Phase IV: U01HG011172 (Cincinnati Children’s Hospital Medical Center); U01HG011175 (Children’s Hospital of Philadelphia); U01HG008680 (Columbia University); U01HG011176 (Icahn School of Medicine at Mount Sinai); U01HG008685 (Mass General Brigham); U01HG006379 (Mayo Clinic); U01HG011169 (Northwestern University); U01HG011167 (University of Alabama at Birmingham); U01HG008657 (University of Washington Medical Center, Seattle); U01HG011181 (Vanderbilt University Medical Center); U01HG011166 (Vanderbilt University Medical Center serving as the Coordinating Center) Phase III: U01HG8657 (Kaiser Permanente Washingtom/University of Washington Medical Center); U01HG8685 (Brigham and Women’s Hospital); U01HG8672 (Vanderbilt University Medical Center); U01HG8666 (Cincinnati Children’s Hospital Medical Center); U01HG6379 (Mayo Clinic); U01HG8679 (Geisinger Clinic); U01HG8680 (Columbia University Health Sciences); U01HG8684 (Children’s Hospital of Philadelphia); U01HG8673 (Northwestern University); U01HG8701 (Vanderbilt University Medical Center serving as the Coordinating Center); U01HG8676 (Partners Healthcare/Broad Institute); and U01HG8664 (Baylor College of Medicine).

