## Supplemental Figures for "Inference of causal relationships based on the genetics of cardiometabolic traits and conditions unique to females in >50,000 participants"

### Supplementary Figure 1: $PRS_{BMI}$ distribution (eMERGE on left and PMBB on right)

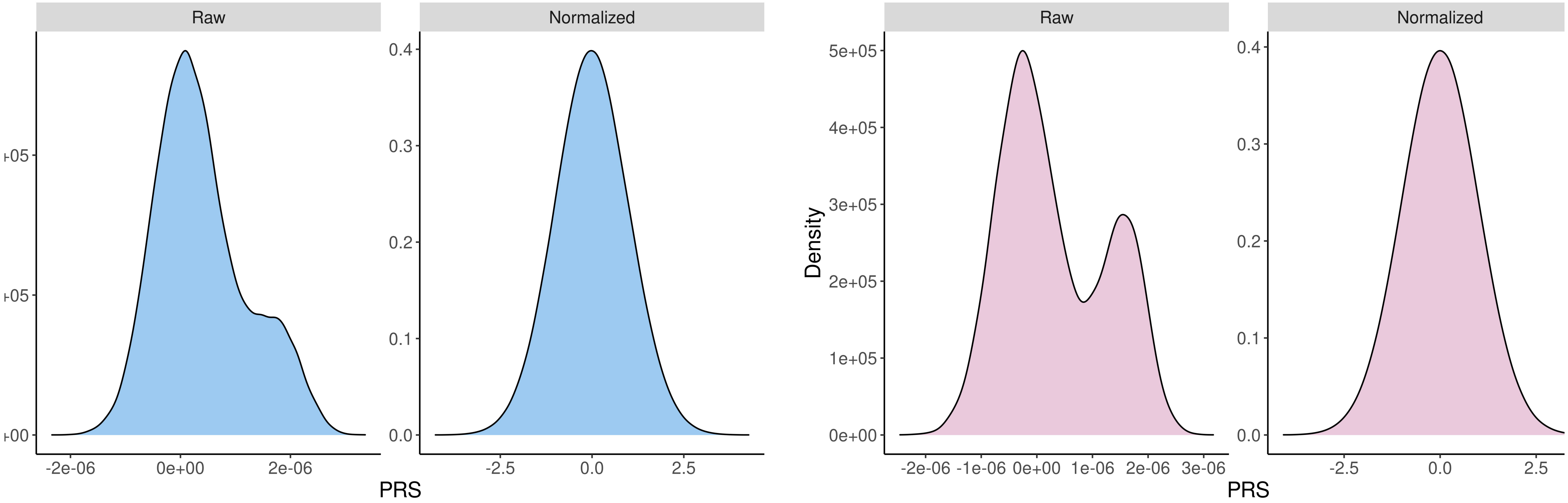

### Supplementary Figure 2: $PRS_{T2D}$ distribution (eMERGE on left and PMBB on right)

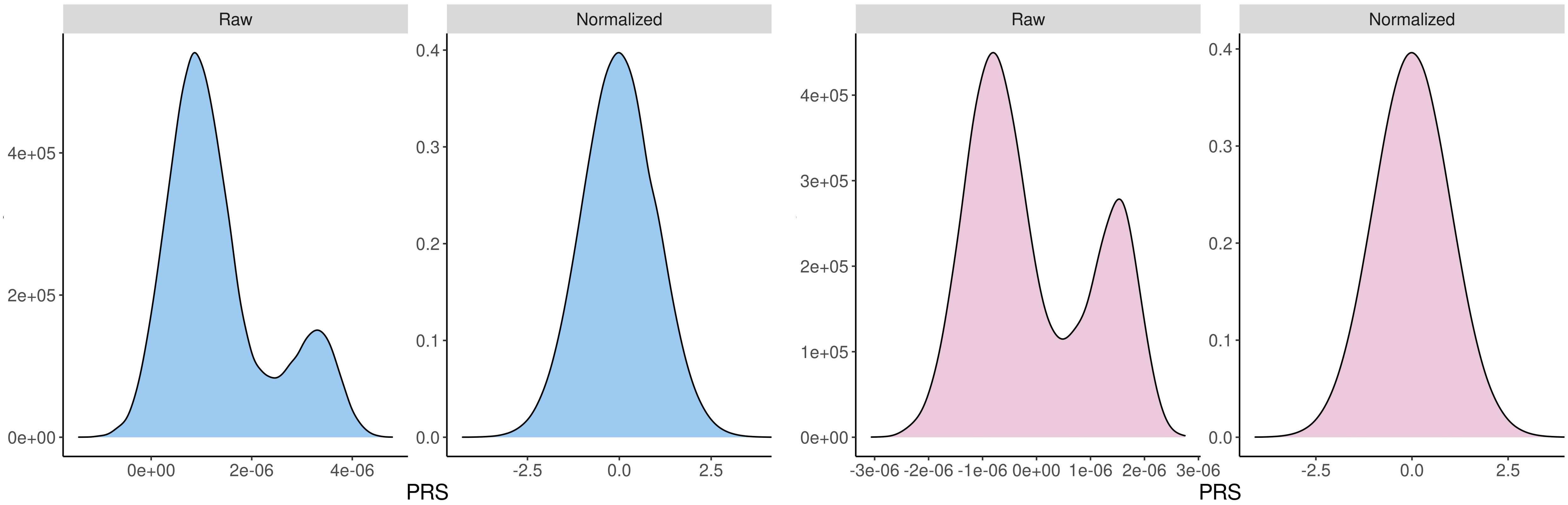

### Supplementary Figure 3: $PRS_{SBP}$ distribution (eMERGE on left and PMBB on right)

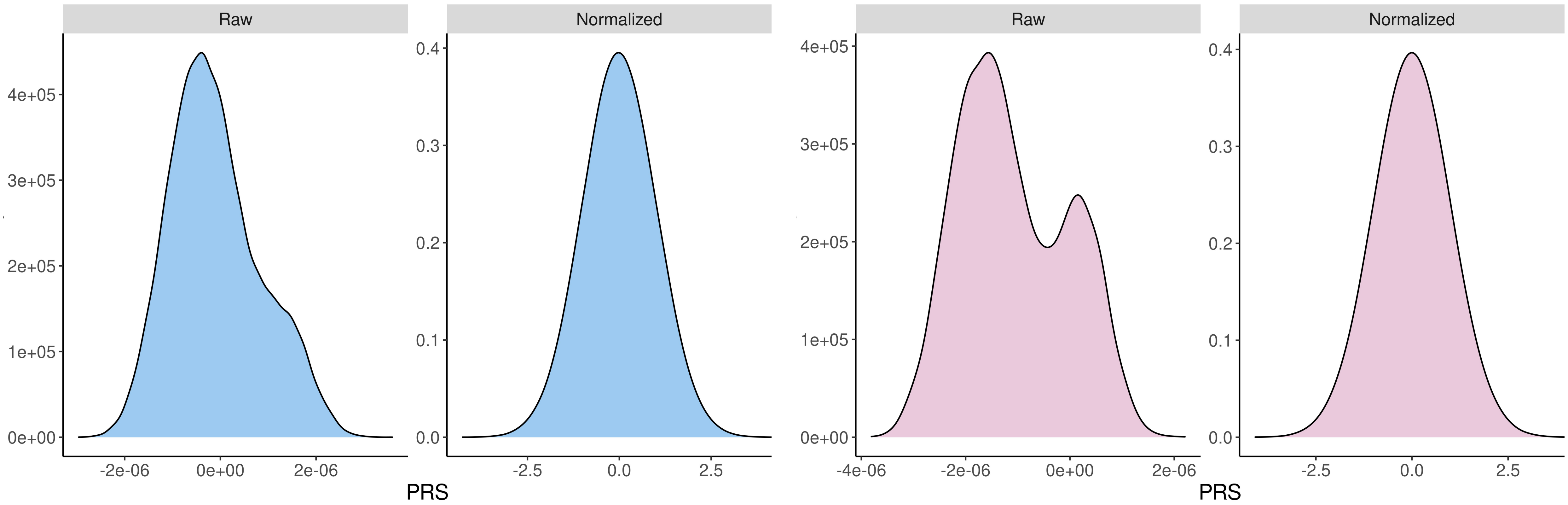

Supplementary Figure 4: $PRS_{DBP}$ distribution (eMERGE on left and PMBB on right)

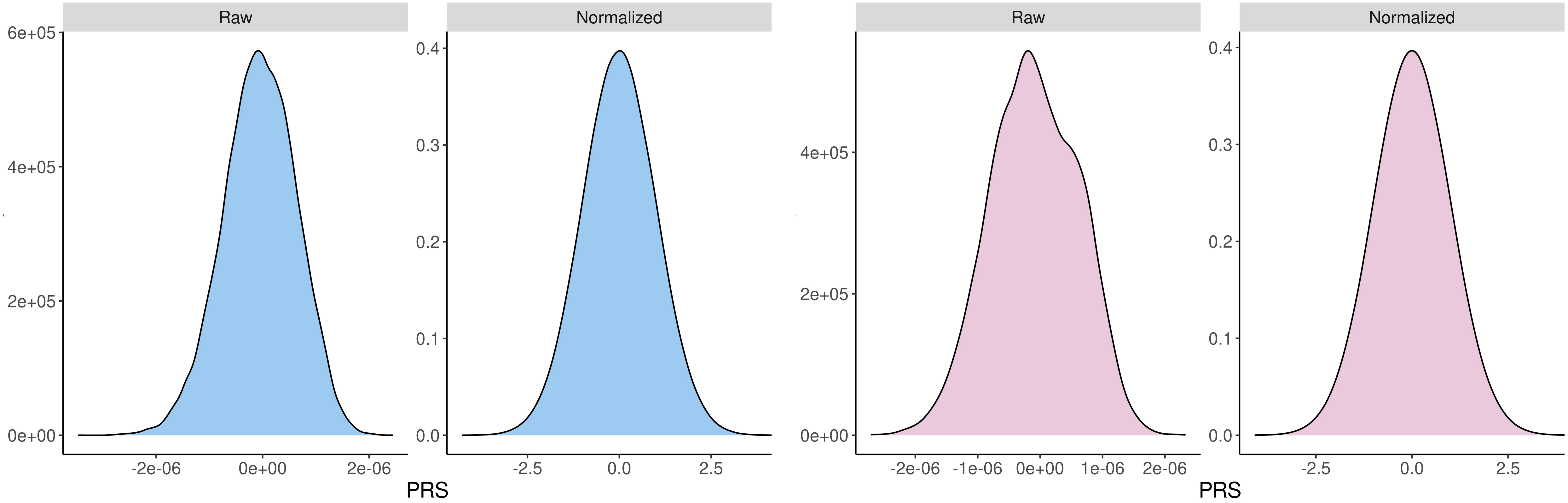

### Supplementary Figure 5: $PRS_{PP}$ distribution (eMERGE on left and PMBB on right)

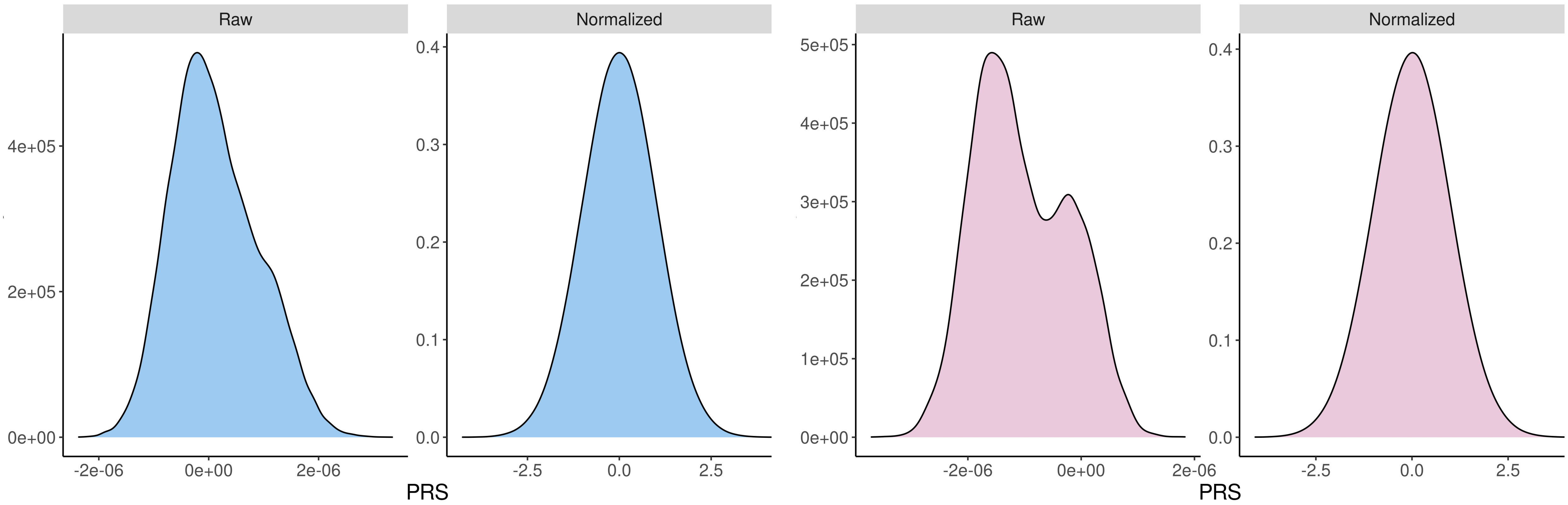

### Supplementary Figure 6: $PRS_{CAD}$ distribution (eMERGE on left and PMBB on right)

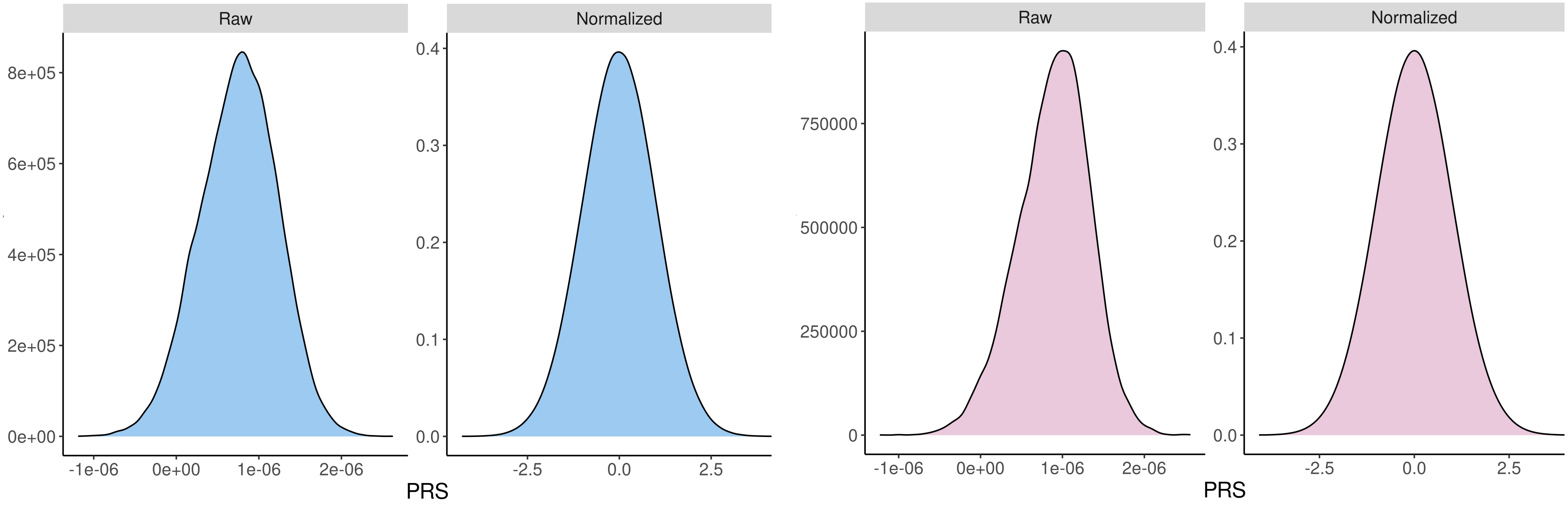

### Supplementary Figure 7: $PRS_{BMI}$ association analyses (eMERGE on right and PMBB on left)

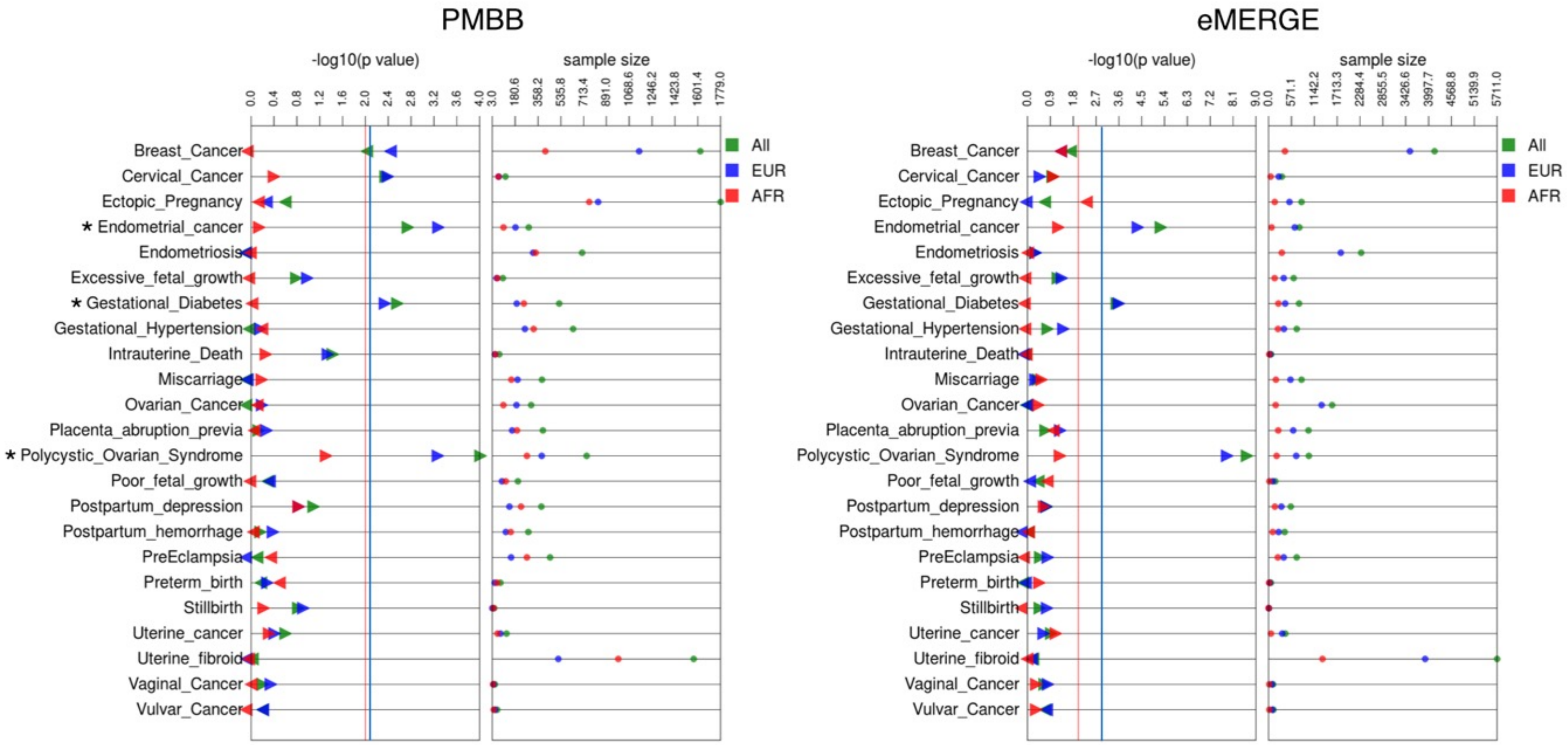

### Supplementary Figure 8: $PRS_{CAD}$ association analyses (eMERGE on right and PMBB on left)

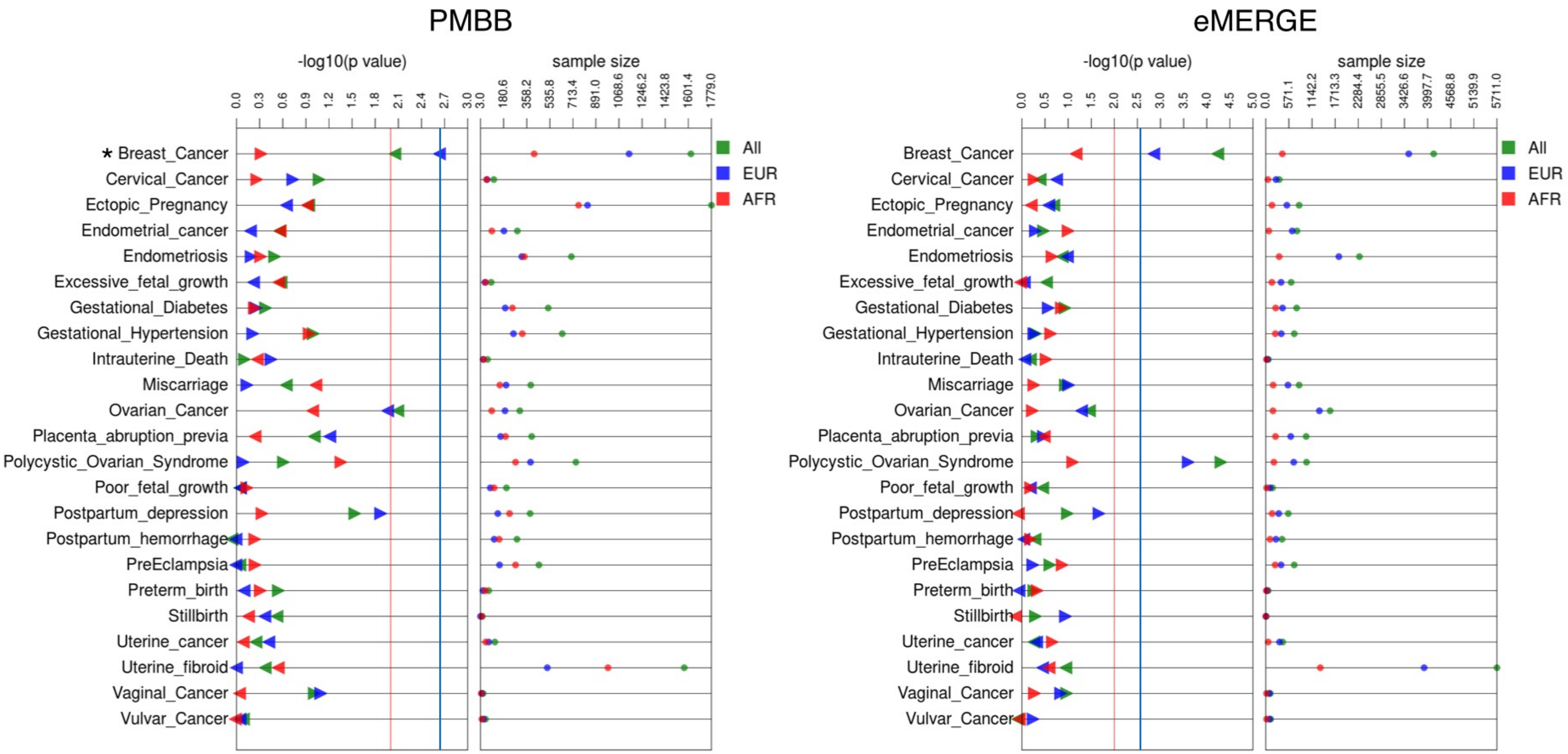

### Supplementary Figure 9: $PRS_{DBP}$ association analyses (eMERGE on right and PMBB on left)

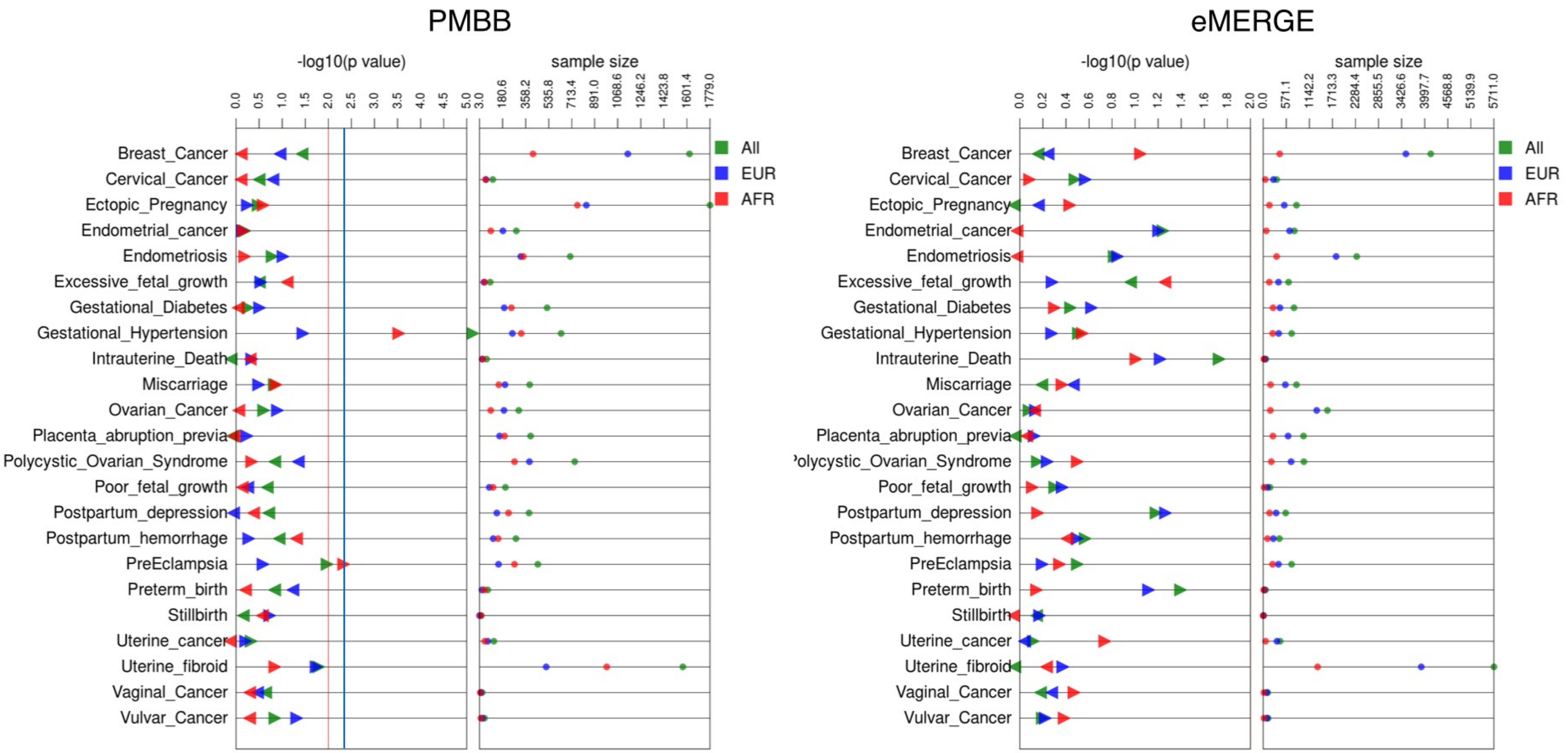

### Supplementary Figure 10: $PRS_{PP}$ association analyses (eMERGE on right and PMBB on left)

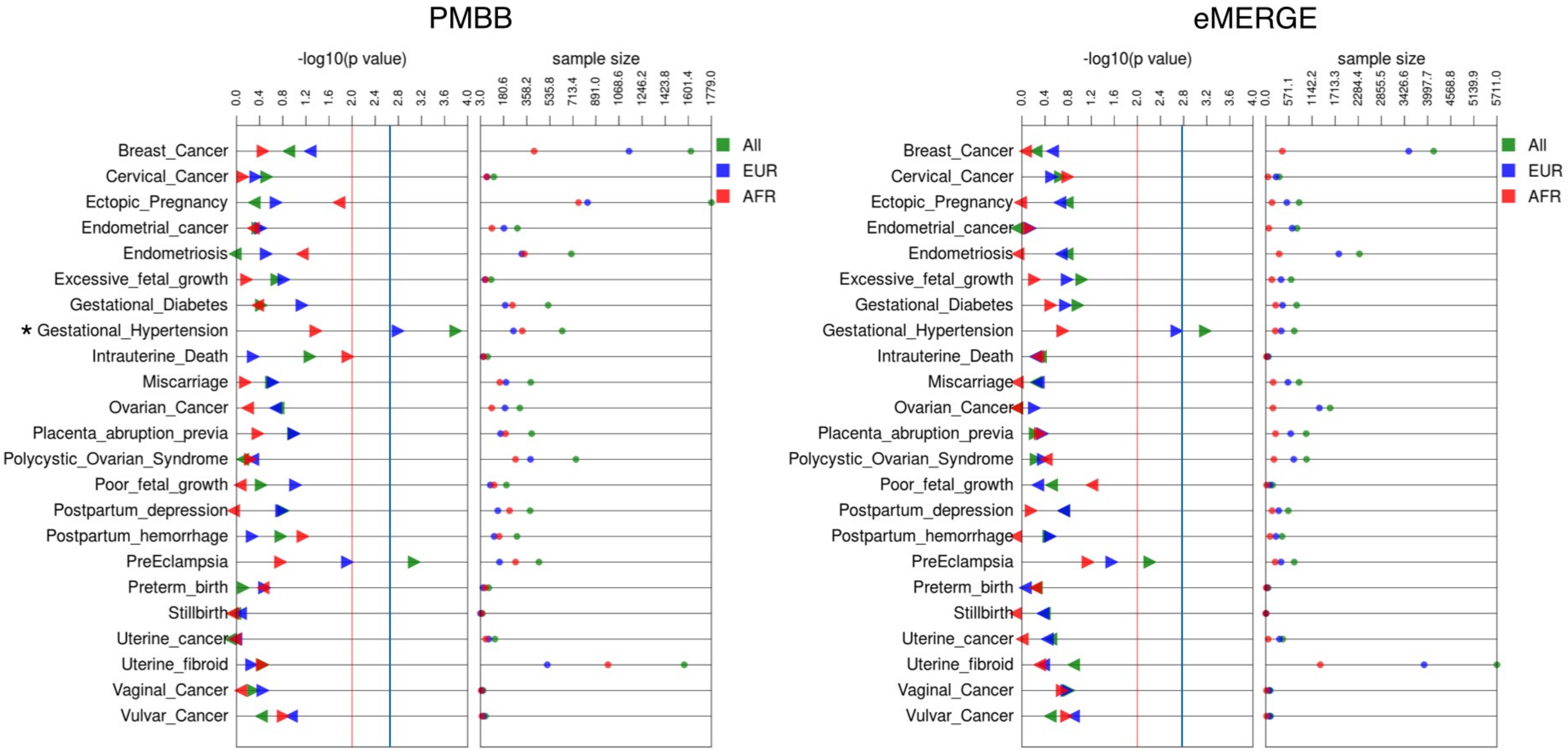

### Supplementary Figure 11: $PRS_{SBP}$ association analyses (eMERGE on right and PMBB on left)

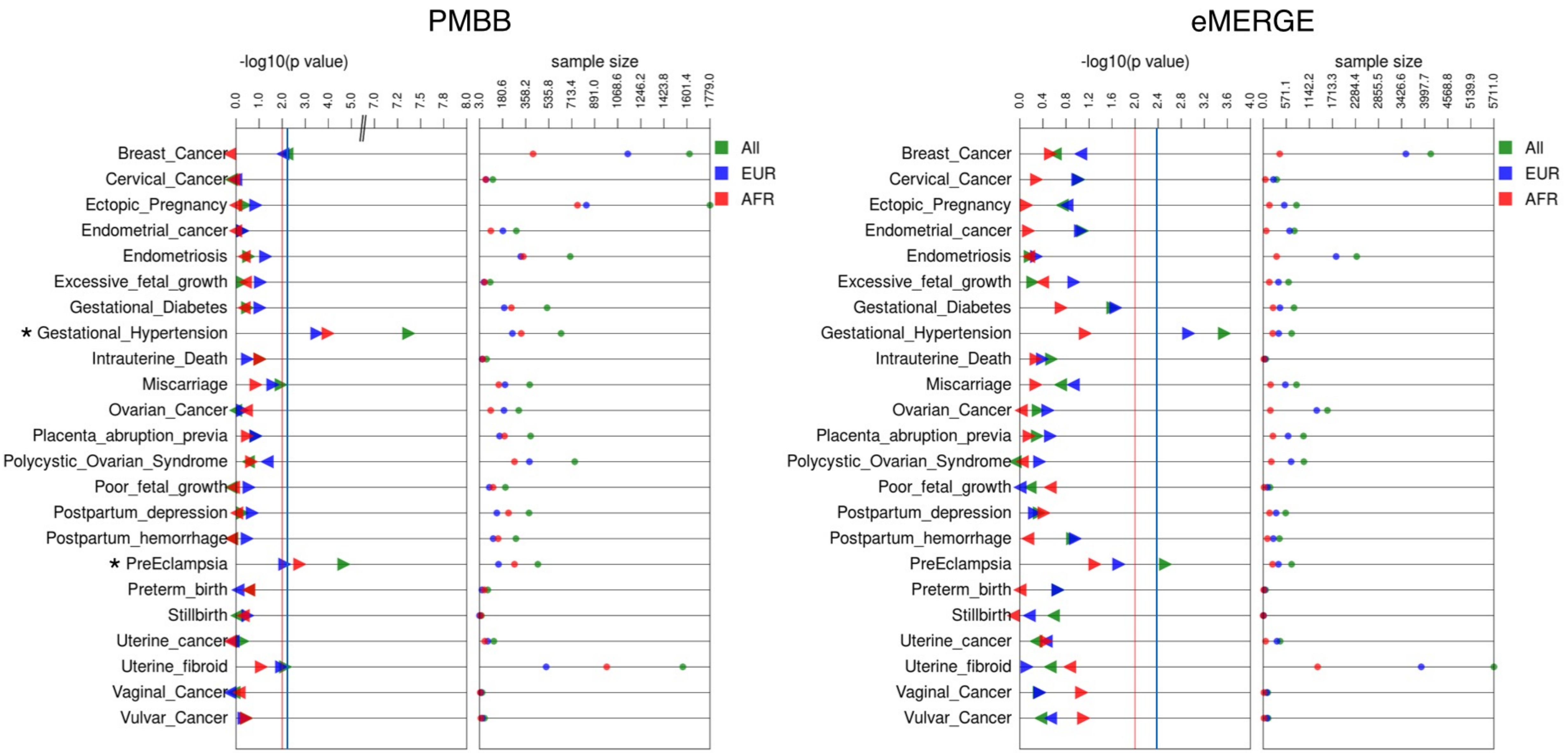

### Supplementary Figure 12: $PRS_{T2D}$ association analyses (eMERGE on right and PMBB on left)

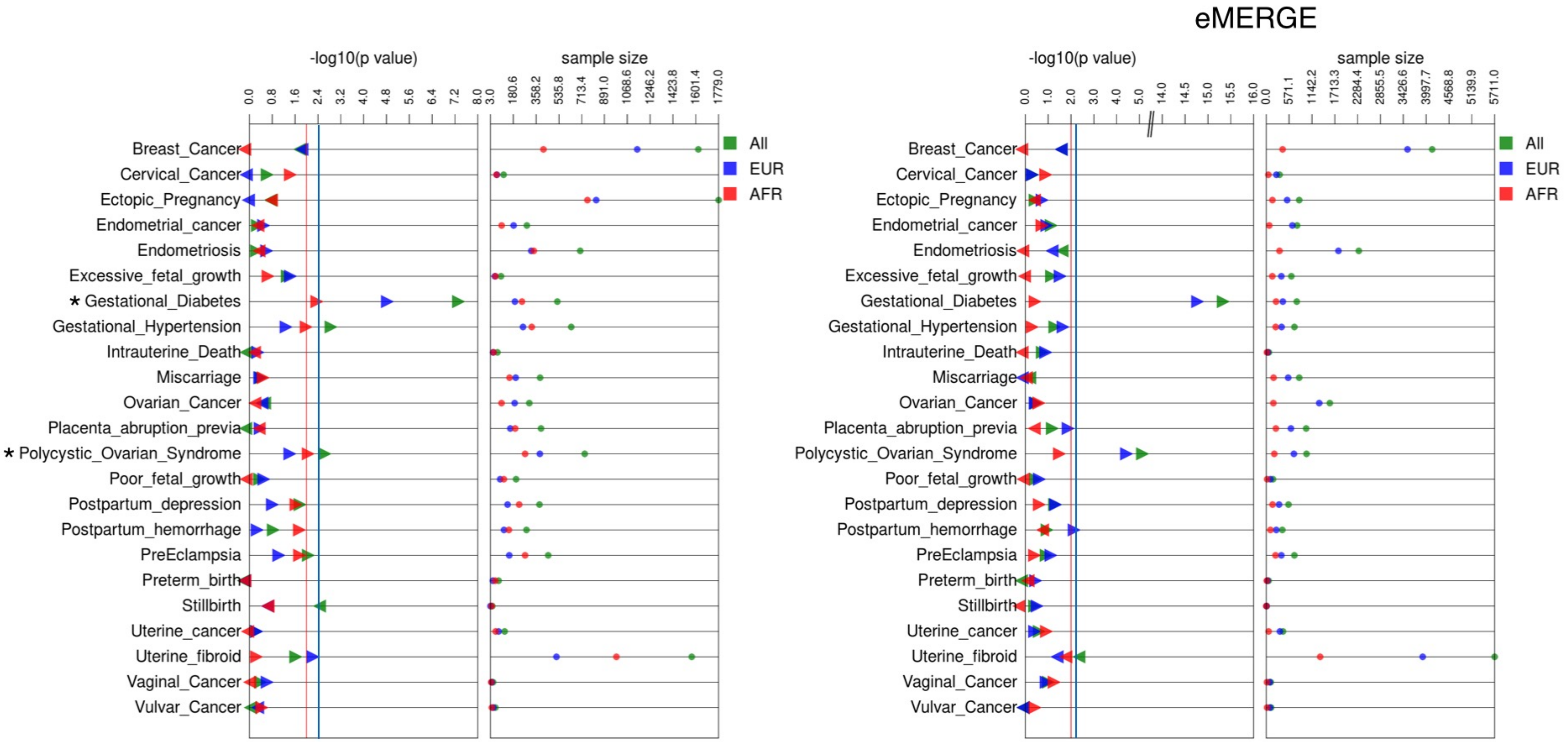

### Supplementary Figure 13: Prevalence by PRS quintile plot (Endometrial cancer and $PRS_{BMI}$ on left and Gestational diabetes and $PRS_{BMI}$ on right)

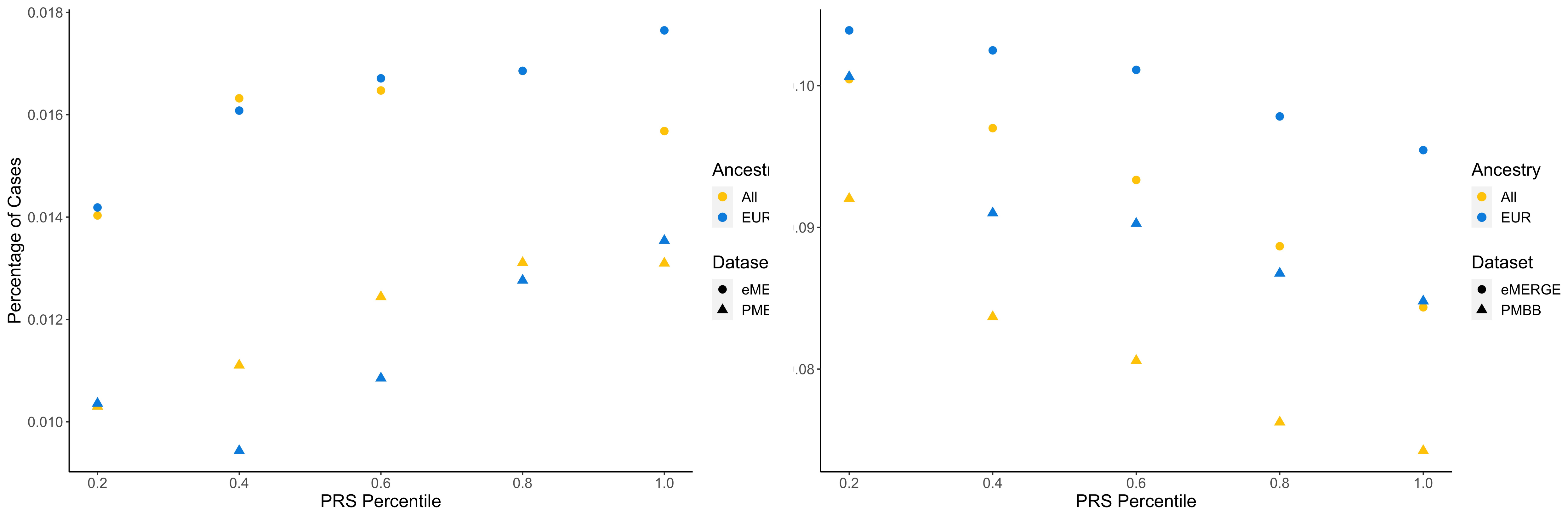

### Supplementary Figure 14: Prevalence by PRS quintile plot (PCOS and $PRS_{BMI}$ on left and Gestational Hypertension and $PRS_{BMI}$ on right)

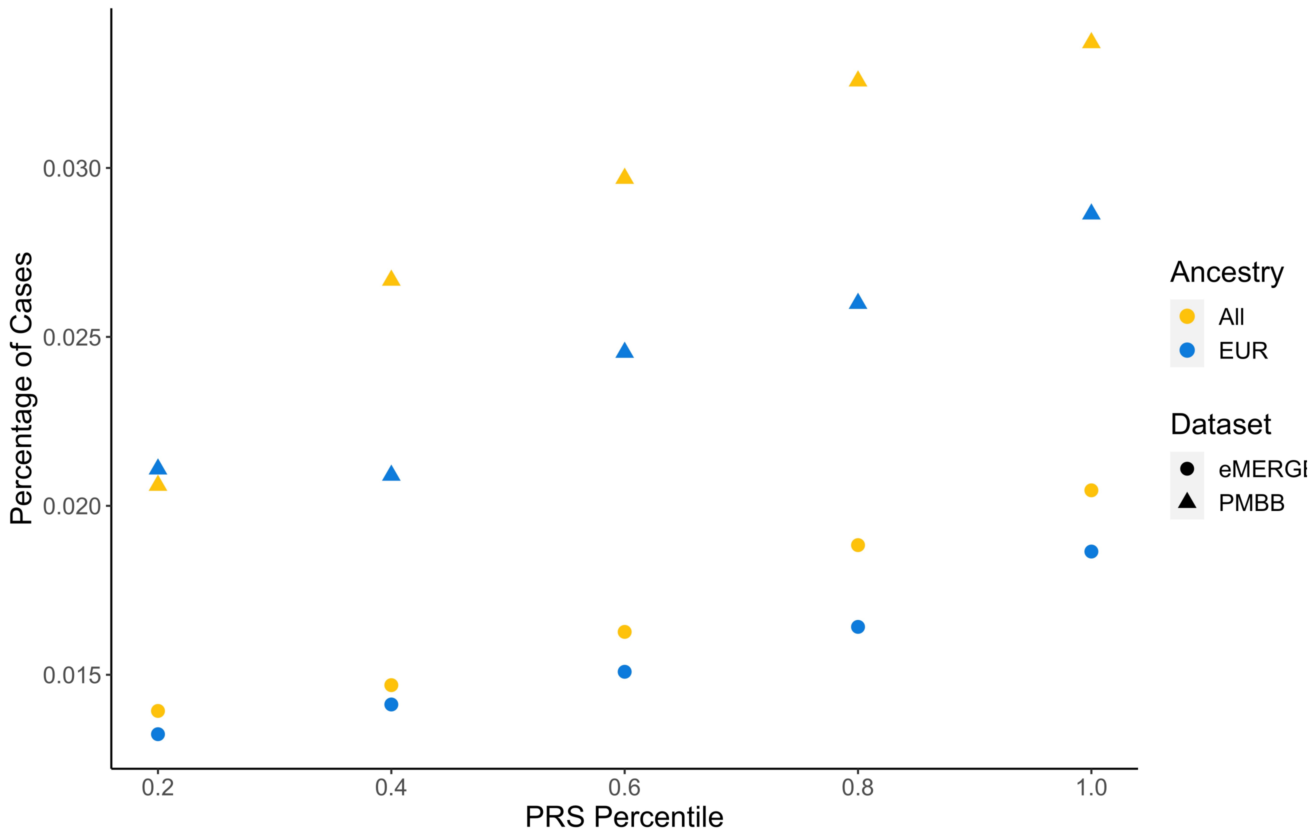


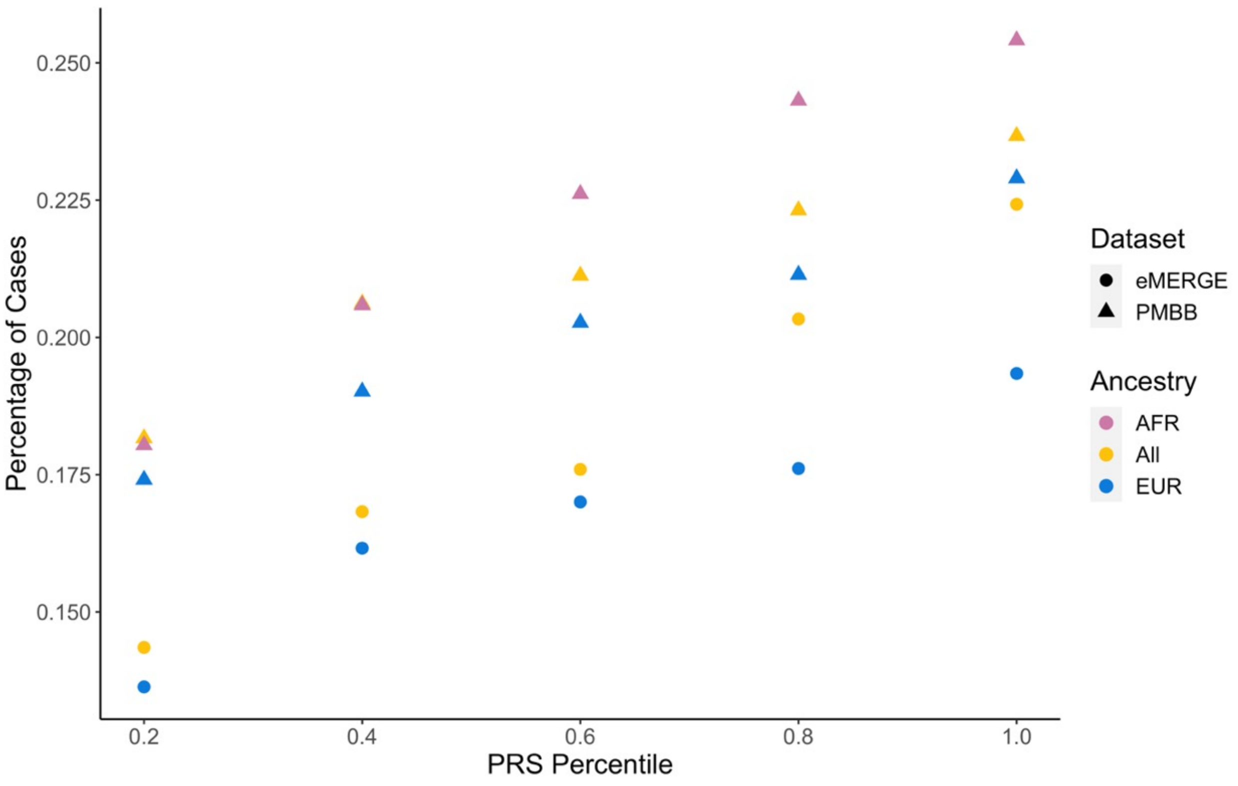

Supplementary Figure 15: Prevalence by PRS quintile plot (Gestational hypertension on left and Preeclampsia (on right) with $PRS_{DBP}$)

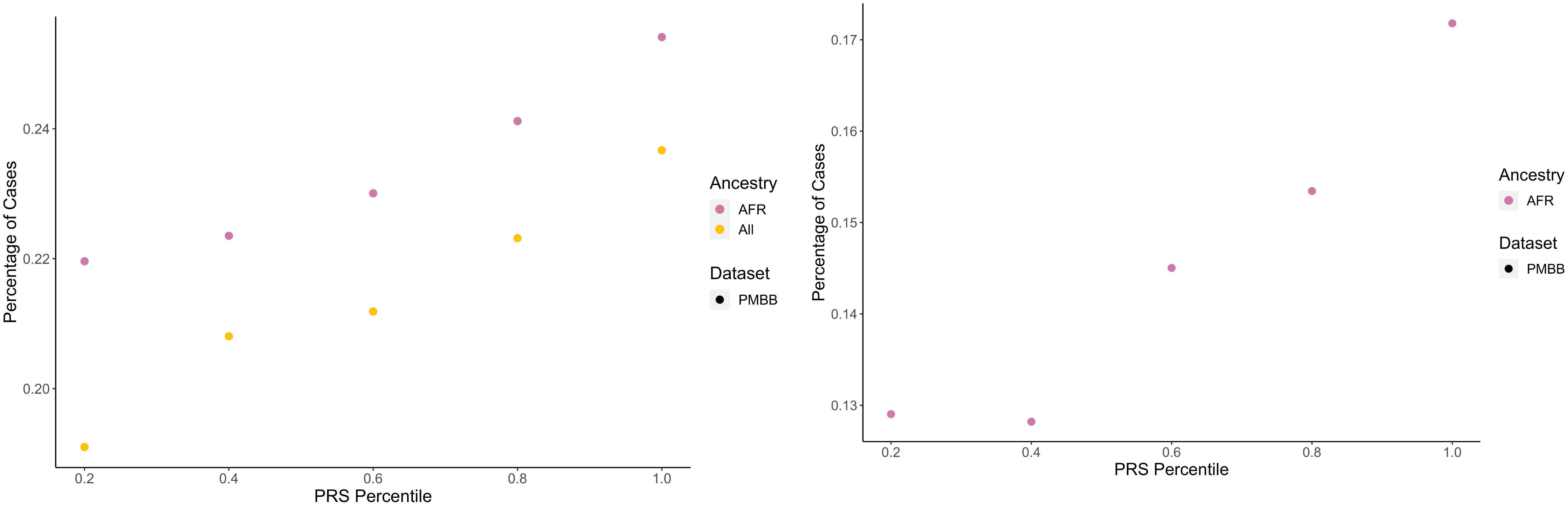

Supplementary Figure 16: Prevalence by PRS quintile plot (Gestational hypertension on left and Preeclampsia (on right) with $PRS_{PP}$)

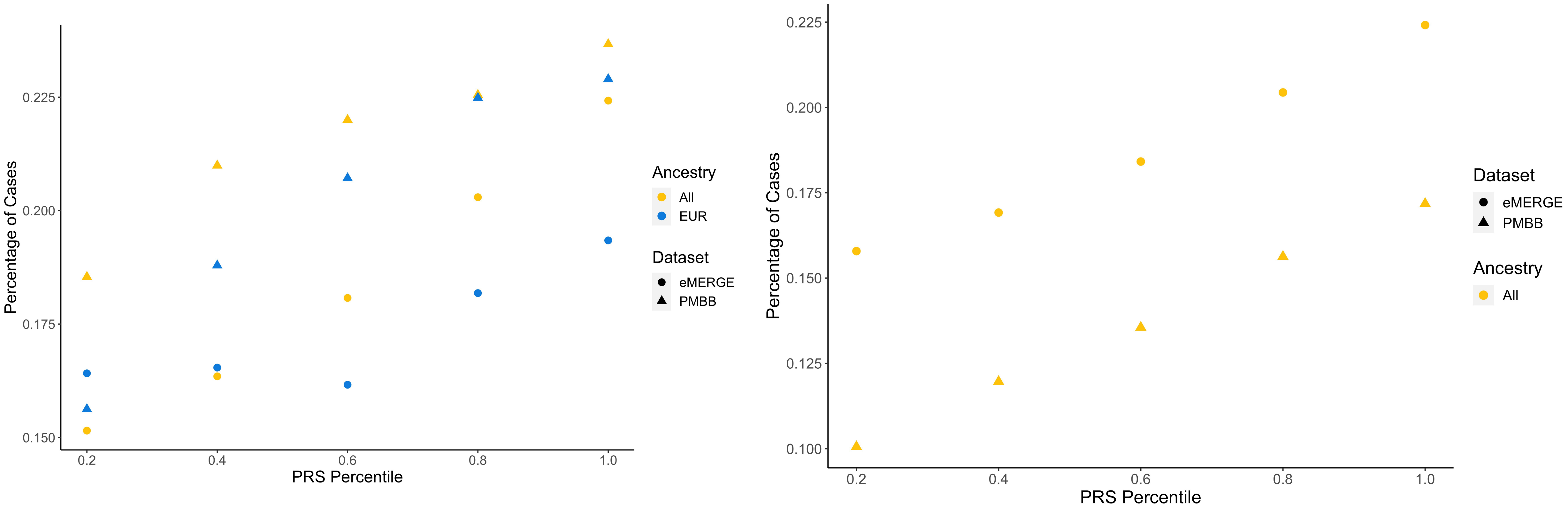

### Supplementary Figure 17: Prevalence by PRS quintile plot (Gestational diabetes on left and PCOS (on right) with $PRS_{T2D}$)

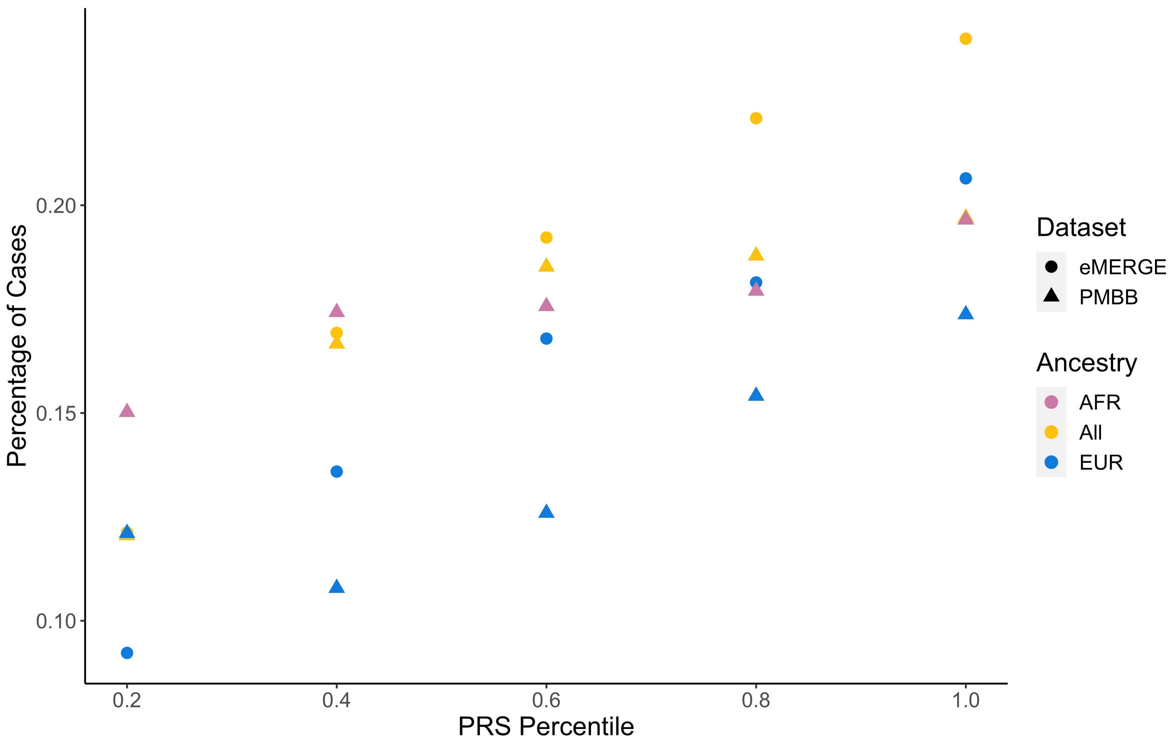


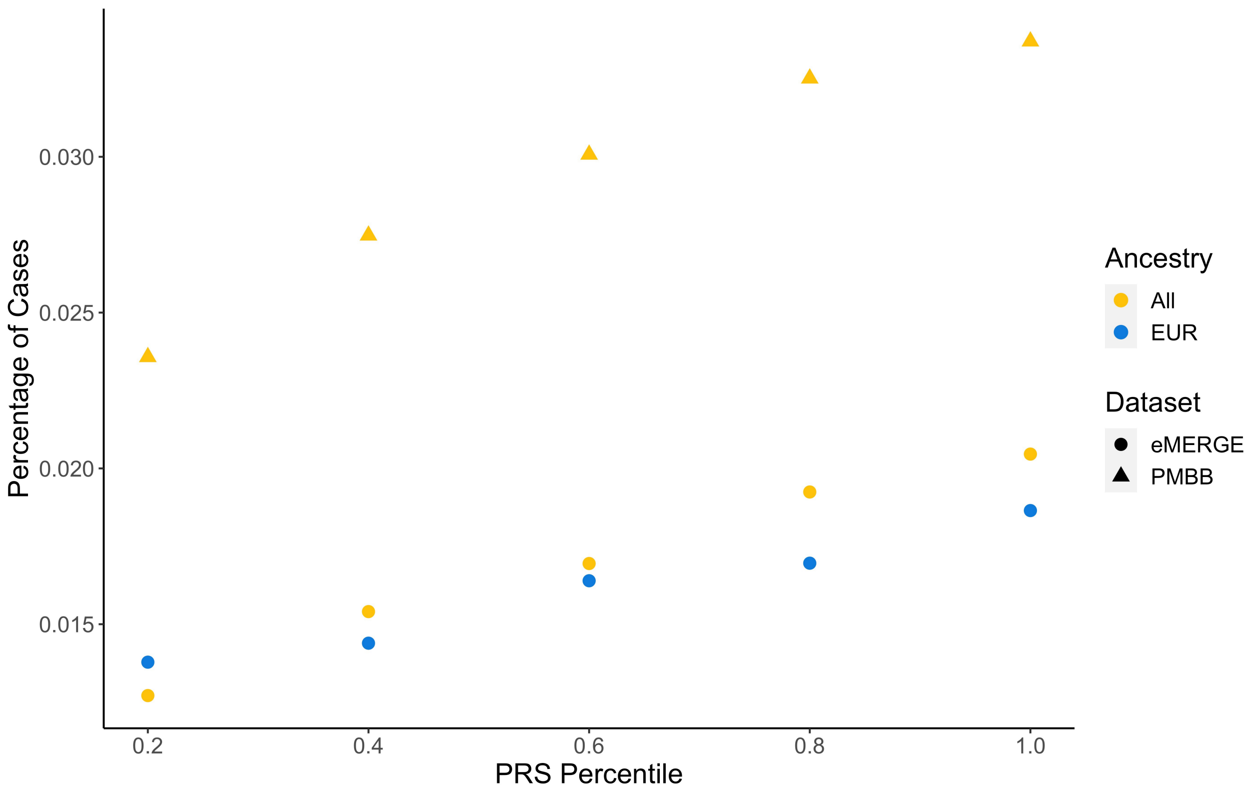

### Supplementary Figure 18: Chronological map for patients with High and Low $PRS_{CAD}$

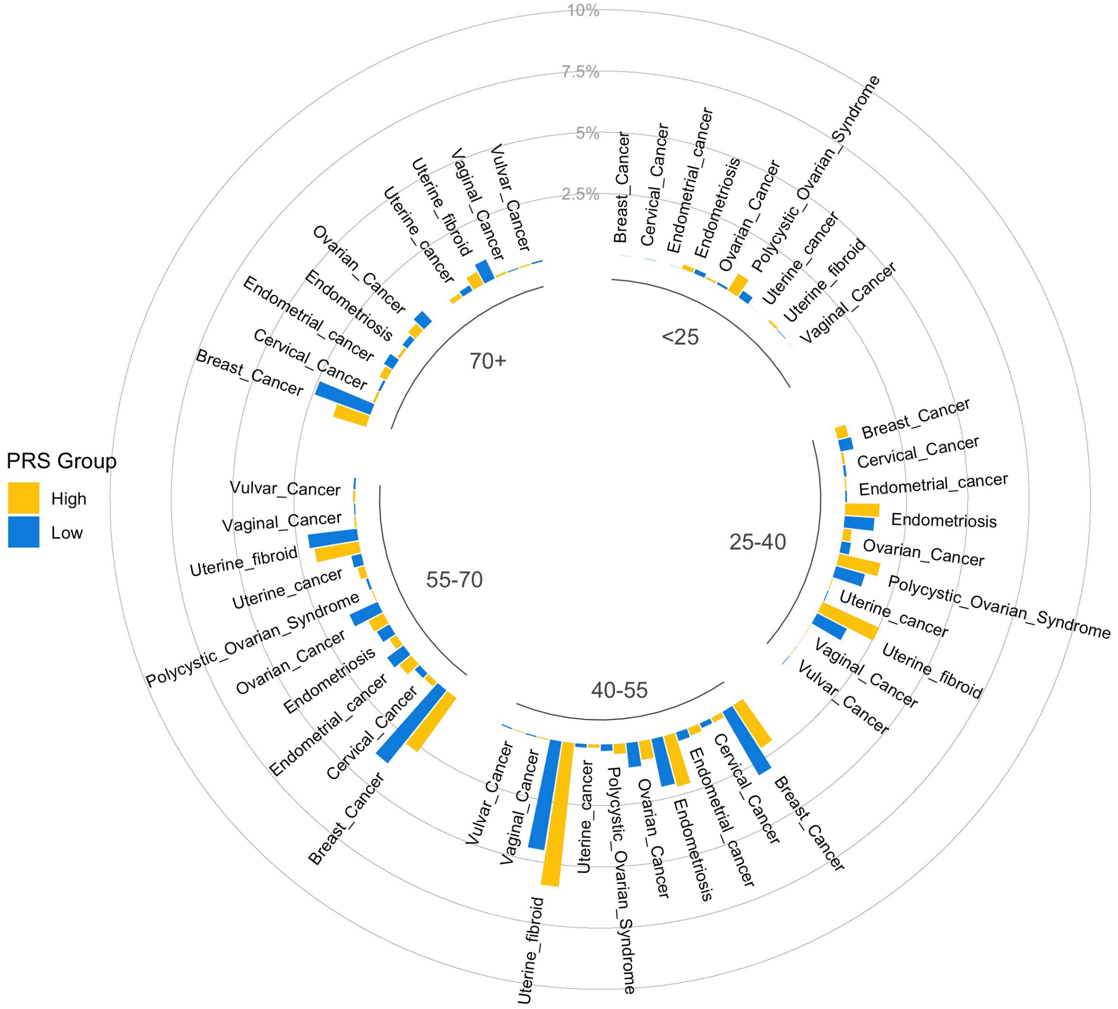


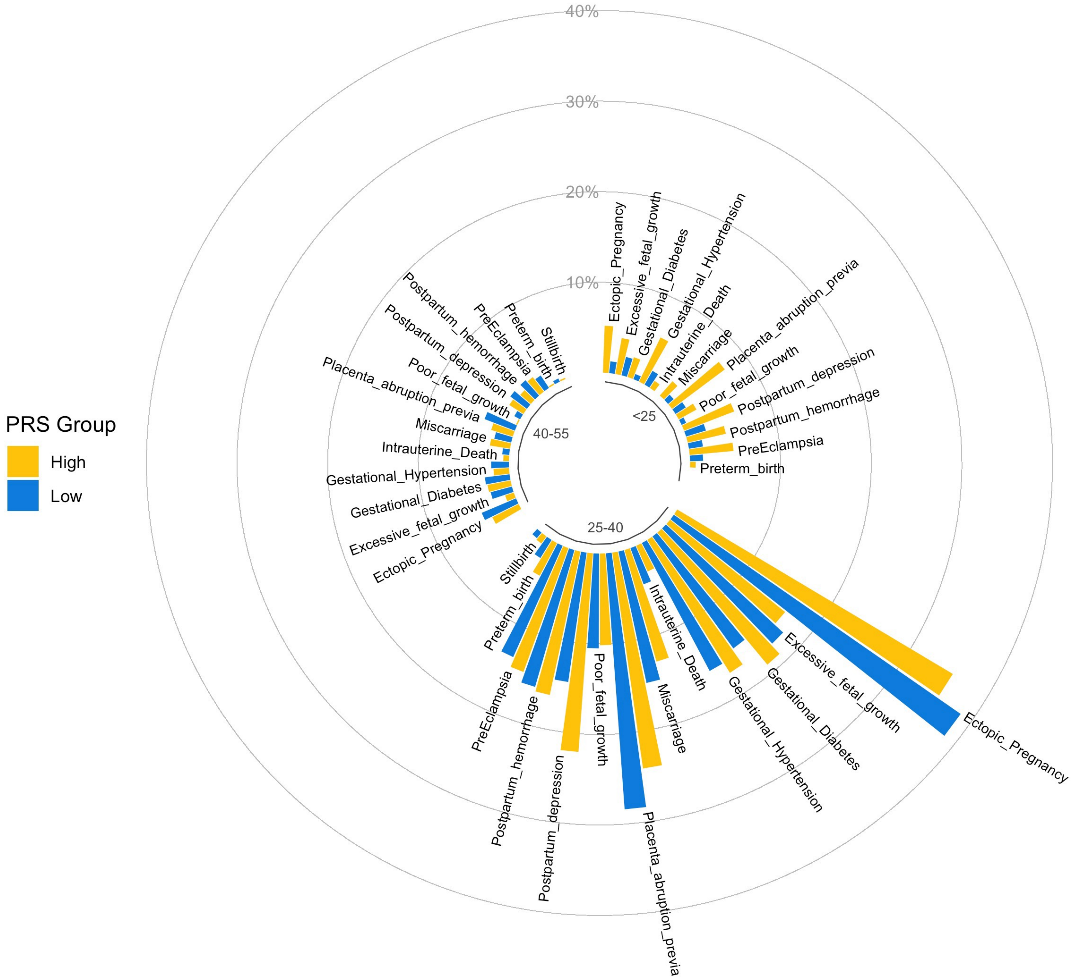

### Supplementary Figure 19: Chronological map for patients with High and Low $PRS_{DBP}$

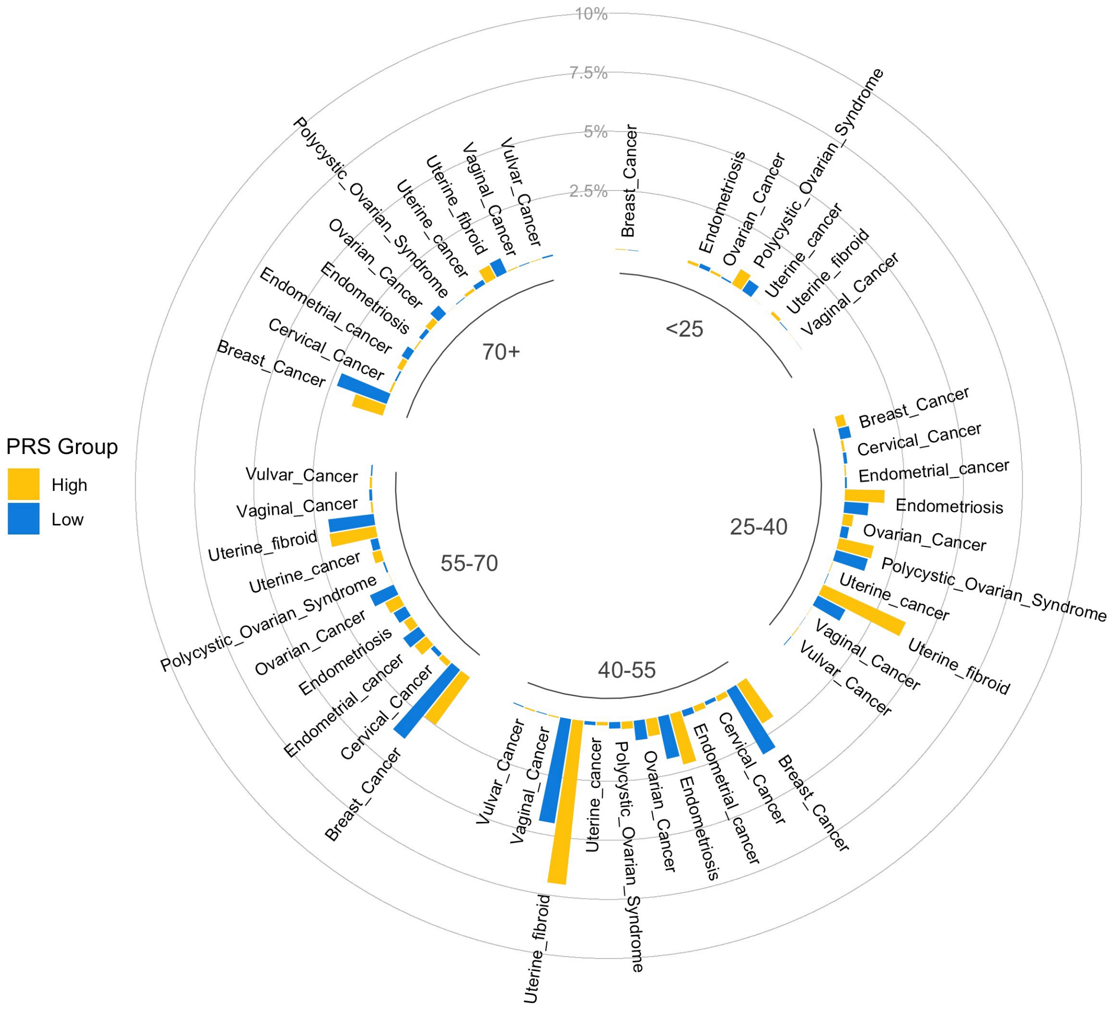


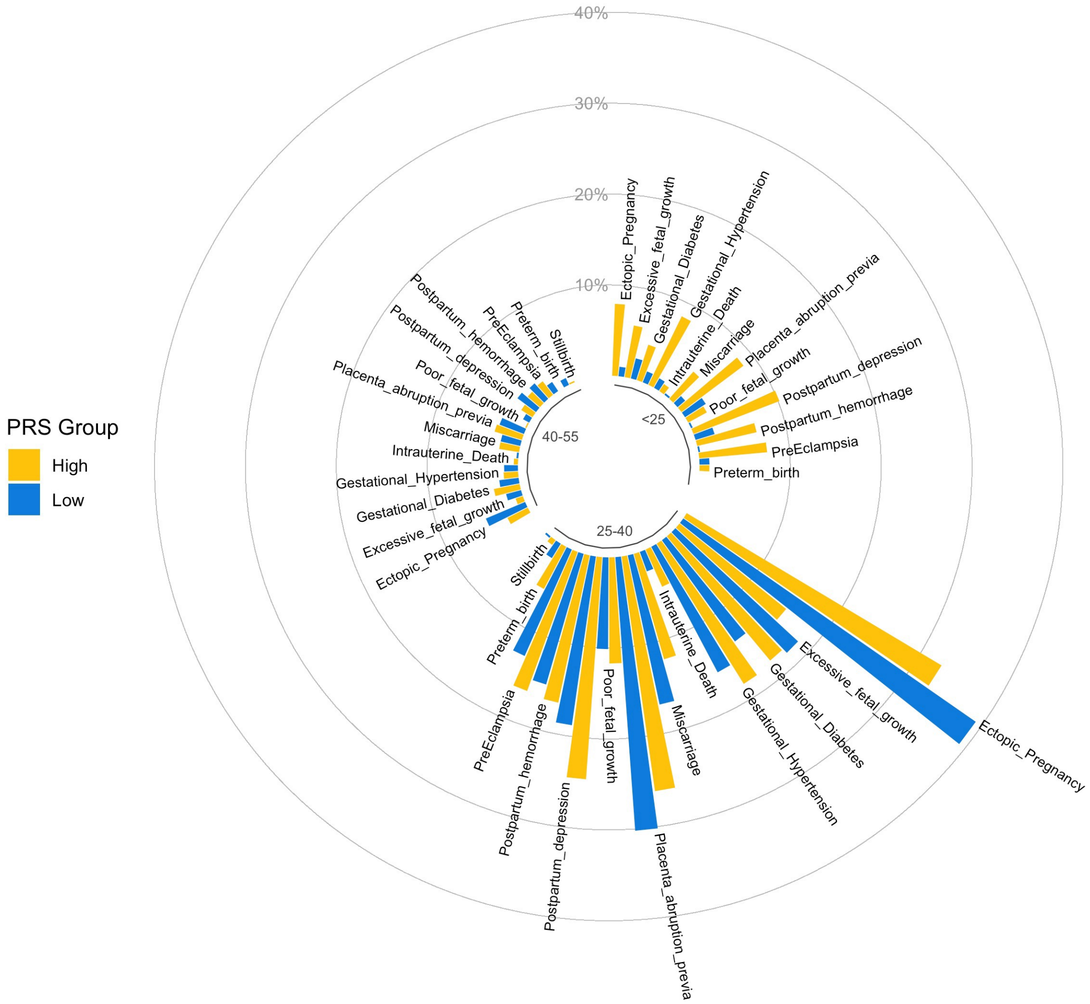

### Supplementary Figure 20: Chronological map for patients with High and Low $PRS_{PP}$

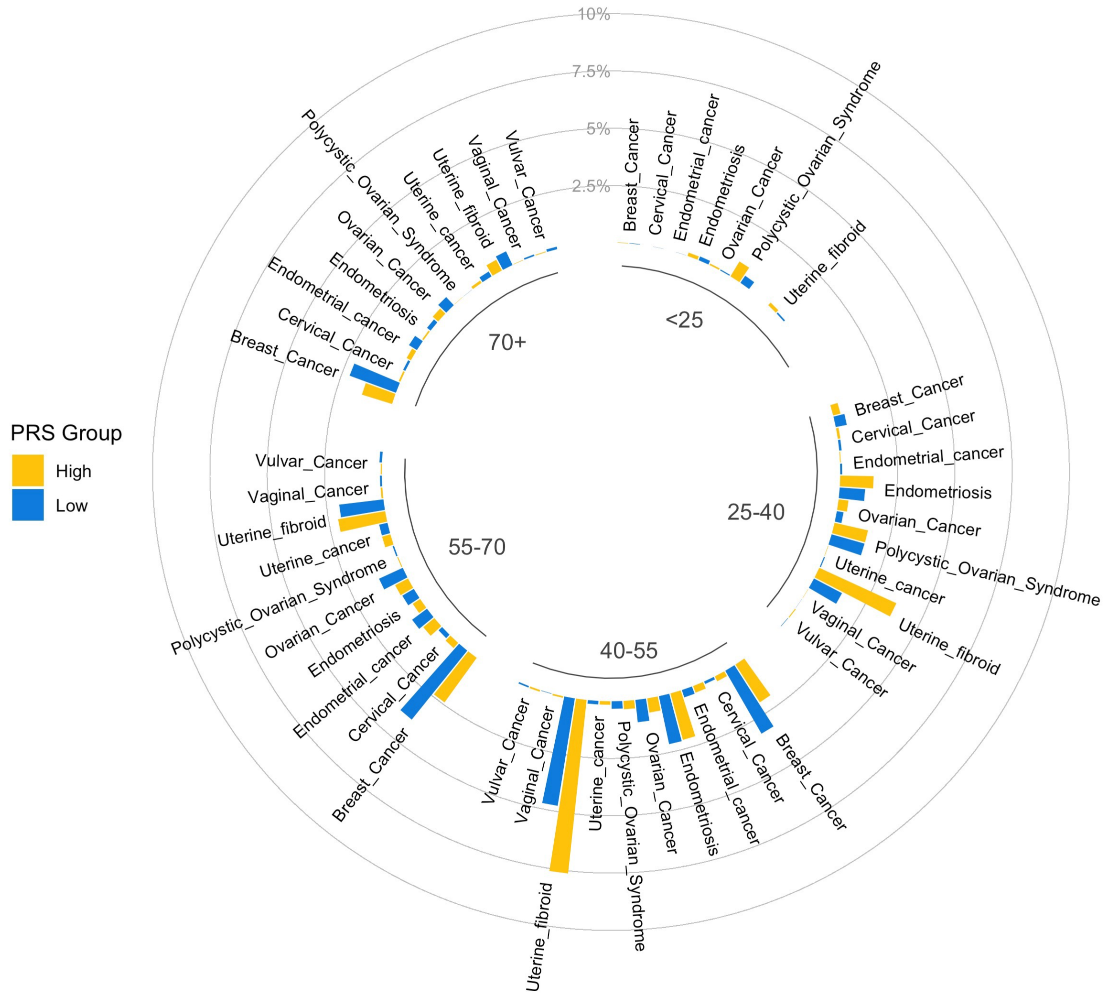


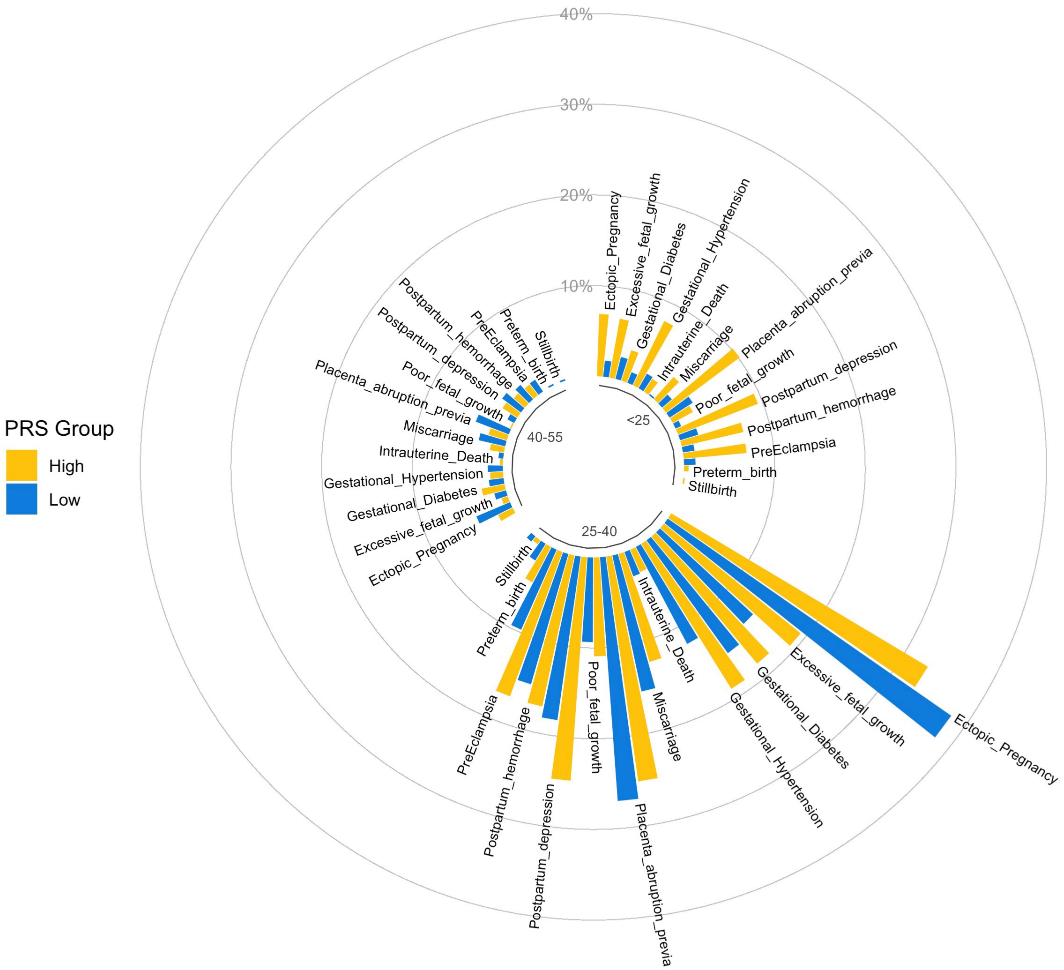

### Supplementary Figure 21: Chronological map for patients with High and Low $PRS_{SBP}$

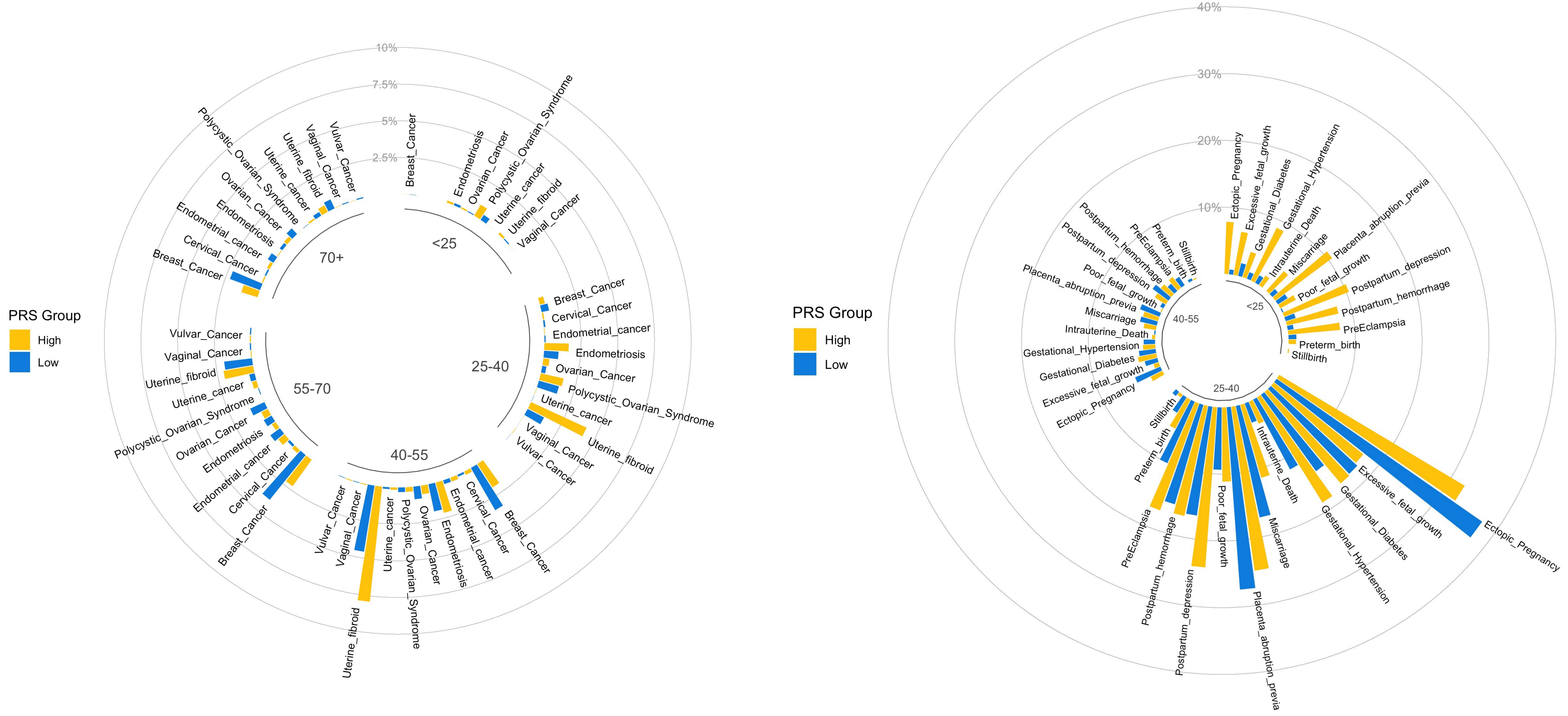

### Supplementary Figure 22: Chronological map for patients with High and Low $PRS_{T2D}$

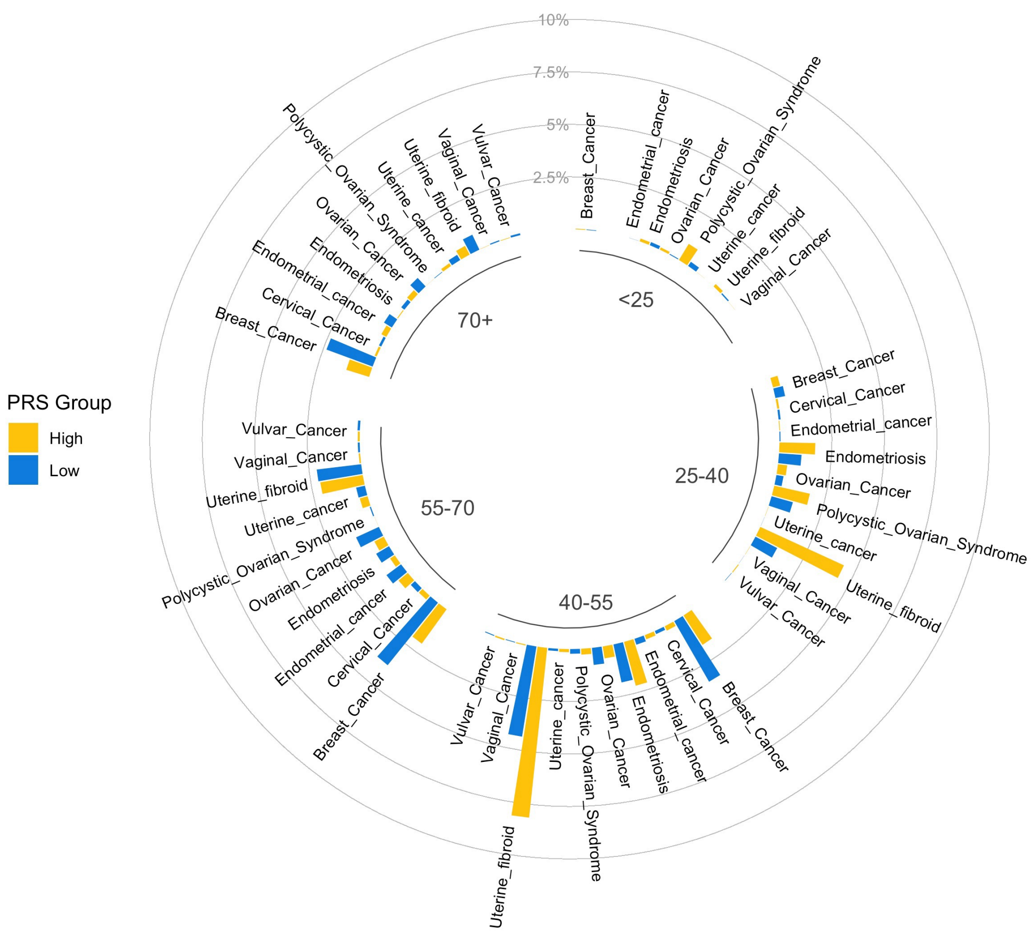


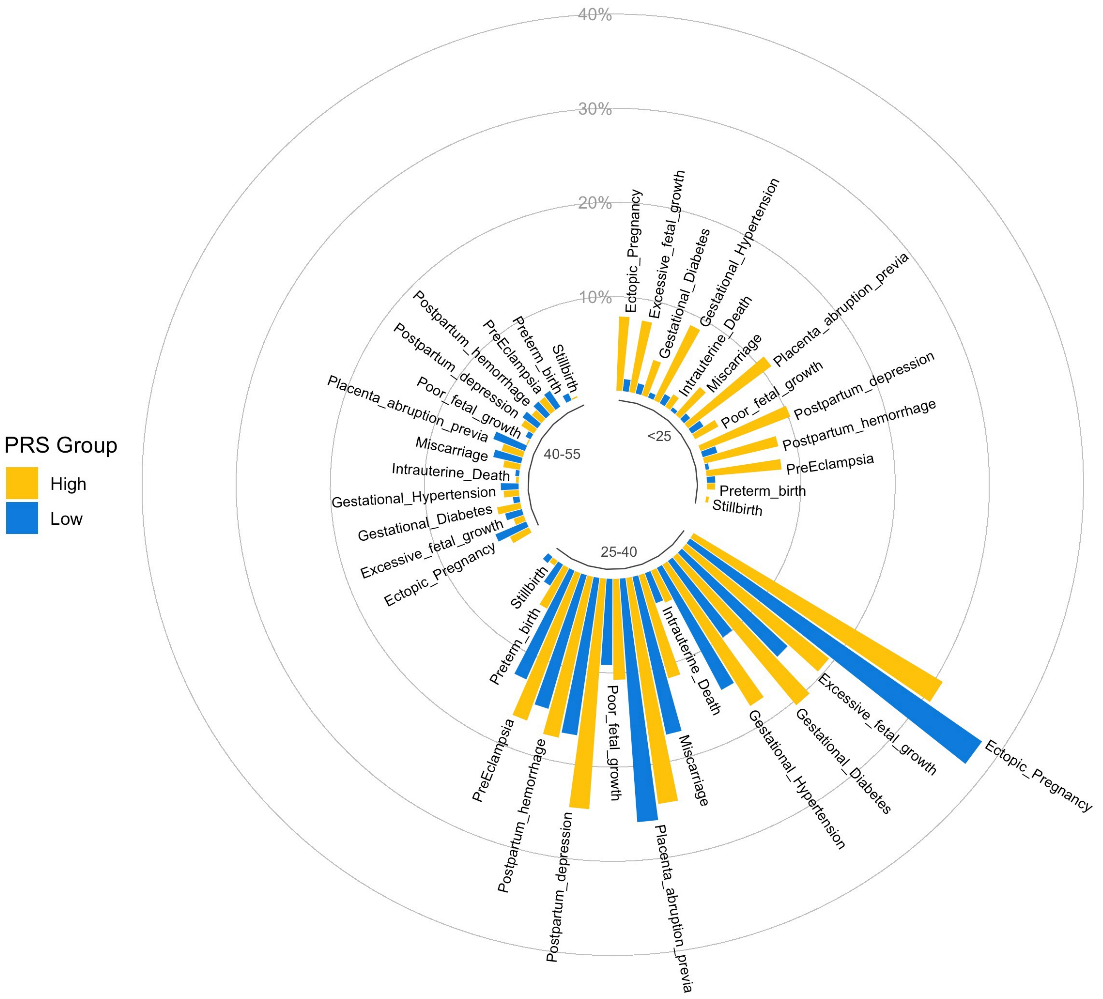
